# The recombinant Link module of human TSG-6 suppresses cartilage damage in models of osteoarthritis: a potential disease-modifying OA drug

**DOI:** 10.1101/2021.03.23.21254102

**Authors:** Sheona P Drummond, Eckart Bartnik, Nikolaos Kouvatsos, Jenny L Scott, Douglas P Dyer, Jennifer M Thomson, Andrew J Price, Sanjay Anand, Leela C Biant, Thomas Leeuw, Matthias Herrmann, Caroline M Milner, Anthony J Day

**Author notes:** ***Correspondence to*:** Professor Anthony J Day, Wellcome Trust Centre for Cell-Matrix Research, Faculty of Biology Medicine & Health, University of Manchester, Michael Smith Building, Oxford Road, Manchester M13 9PT, UK;, Dr Caroline M Milner, Faculty of Biology Medicine & Health, University of Manchester, Michael Smith Building, Oxford Road, Manchester M13 9PT, UK.

## Abstract

**Objectives:** To investigate the role of endogenous TSG-6 in human osteoarthritis (OA) and assess the disease-modifying potential of a TSG-6-based biological treatment in cell, explant and animal models of OA.

**Methods:** Knee articular cartilages from OA patients were analysed for TSG-6 protein and mRNA expression using immunohistochemistry and RNAscope, respectively. The inhibitory activities of TSG-6 and its isolated Link module domain (Link_TSG6) on cytokine-induced glycosaminoglycan loss in OA cartilage explants were compared. Mesenchymal stem/stromal cell (MSC)-derived chondrocyte pellet cultures were used to determine the effects of Link_TSG6 and full-length TSG-6 on IL-1α-, IL-1β- or TNF-stimulated *ADAMTS4, ADAMTS5* and *MMP13* mRNA expression. Link_TSG6 was administered *i.a*. to the rat ACLTpMMx model and cartilage damage and tactile allodynia were assayed.

**Results:** TSG-6 is predominantly associated with chondrocytes in regions of cartilage damage and its expression is negatively correlated with MMP13, the major collagenase implicated in OA progression. Link_TSG6 is more potent than full-length TSG-6 at dose-dependently inhibiting cytokine-mediated matrix breakdown in human OA cartilage explants; about 50% of donor cartilages, from 59 tested, were responsive to Link_TSG6 treatment. Similarly, Link_TSG6 displayed more potent effects in 3D pellet cultures, suppressing aggrecanase and collagenase gene expression. Link_TSG6 treatment reduced touch-evoked pain and dose-dependently inhibited cartilage damage in a rodent model of surgically-induced OA.

**Conclusions:** Native TSG-6 is associated with a low catabolic chondrocyte phenotype in OA cartilage. Link_TSG6, which has enhanced chondroprotective activity compared to the full-length TSG-6 protein, demonstrates potential as a disease modifying OA drug (DMOAD) and warrants further investigation and development.

**KEY MESSAGES:** *What is already known about this subject?:*

- TSG-6, a protein with anti-inflammatory and protective effects in other tissues, is expressed in joints affected by osteoarthritis – a condition for which there are no disease-modifying drugs.

*What does this study add?:*

- A novel protective mechanism has been identified, whereby TSG-6 inhibits inflammatory cytokine-induced catabolic pathways in cartilage.
- The Link module of TSG-6 (Link_TSG6) has been shown to have greater potency than TSG-6 as an inhibitor of cartilage damage and is efficacious in a rat model of osteoarthritis.
- Data from cartilage explants indicate that OA patients can be stratified for Link_TSG6 responsiveness.

*How might this impact on clinical practice or future developments?:*

- Link_TSG6 has been identified as potential disease-modifying OA drug that mimics an intrinsic protective process.

## INTRODUCTION

Osteoarthritis (OA) is the most common form of joint disease, where associated pain and functional decline represent a major and growing cause of long-term disability[1-3]. A large proportion of this is accounted for by knee OA, which is estimated to affect >250 million people worldwide[3]. OA is a complex disorder where constitutional (e.g. age, obesity), biomechanical (e.g. joint injury, occupational activity) and genetic factors contribute to structural deterioration and, potentially, failure of the synovial joint (reviewed in[4,5]). Whilst degeneration of the cartilage matrix is a major hallmark of OA, the whole joint is affected; for example, synovitis and pathological changes to subchondral bone are common features of the disease. At present there are no licensed disease-modifying OA drugs (DMOAD) and strategies for pain relief are inadequate[6]. Ultimately, a large number of patients undergo joint arthroplasty for late stage OA[7,8] with outcomes being unsatisfactory in a significant proportion of cases[9,10].

Joint injury and obesity are major risk factors for knee OA, consistent with the contribution of adverse mechanical loading to disease onset and progression. What is not clear is why some individuals experience progressive, symptomatic OA while others with similar structural changes to the joint do not (see[11]). The balance between degenerative and protective mechanisms in cartilage is likely an important factor and these processes provide potential targets for new therapeutic strategies. The principle catabolic enzymes that drive cartilage degradation during OA are the collagenase MMP13 and the ADAMTS family of aggrecanases, particularly ADAMTS4 and ADAMTS5[12,13]. These proteases are expressed by chondrocytes and mediate the loss of the proteoglycan aggrecan that is a pre-requisite for irreversible breakdown of type II collagen. Animal models where joints are surgically destabilised have been validated as recapitulating many aspects of the OA disease process (see[14]). Studies in such models have identified that the immediate response to injury occurs primarily in the cartilage, is inflammatory in nature and involves elevated expression of both pro- and anti-catabolic factors[15,16]. For example genes encoding cytokines (*e*.*g. Il1*), *Adamts4* and *Adamts5*, are rapidly up-regulated following joint destabilisation in mice; expression of *Tsg6*, which is associated with protective/reparative effects in other tissues, is also increased[15].

*TSG-6* gene expression is induced by inflammatory cytokines, for example in IL-1-treated chondrocytes from OA patients[17]. Recent analyses of chondrocyte transcriptomes have consistently identified *TSG-6* (also known as *TNFAIP6*) as one of the most highly up-regulated genes in damaged regions of OA cartilage compared to intact tissue from the same joint[18-20] Furthermore, TSG-6 is secreted by mesenchymal stem/stromal cells (MSC) in response to inflammatory signals and mediates tissue protective and immunomodulatory effects in a diversity of disease models (reviewed in[21,22]). Several mechanisms of action have been described for these activities. For example, we have shown that TSG-6 (and its isolated Link module domain; Link_TSG6) inhibits neutrophil migration[23] via interaction with the chemokine CXCL8[24]. This anti-inflammatory activity of TSG-6 was suggested to underpin its chondroprotective effects in models of rheumatoid arthritis (RA), where *Tsg6-*null mice develop much more severe joint damage with extensive neutrophil infiltration[25]. Treatment with TSG-6 protein or its overexpression in RA models were found to significantly reduce joint damage[26-28]. Moreover, TSG-6 can suppress inflammation by other mechanisms; for example, via down regulation of NF-ĸB-, JNK- and p38-mediated signalling pathways, reducing cytokine expression and promoting polarisation of macrophages towards an M2 (anti-inflammatory) phenotype (see[22]).

The local expression of TSG-6 in OA joints[18-20,29] and its protective effects in RA suggest that this protein is part of an intrinsic repair pathway that could be targeted in the treatment of OA. Conversely, it has been hypothesized that TSG-6 contributes to the OA disease process[30]. There are no data on the direct effects of TSG-6 on chondrocyte activity and there has been little exploration of its disease-modulating effects in models of OA.

## METHODS

See online supplemental material.

## RESULTS

### TSG-6 protein is associated with damaged regions of OA cartilage

TSG-6 is expressed by chondrocytes in response to inflammatory stimuli[17,31] and both cell- and matrix-associated protein has been detected in OA cartilage[29,30]. Here, we analysed cartilage from patients with anteromedial gonarthrosis (AMG; Figure 1A), where the medial tibial plateau displays a characteristic pattern of erosion (Supplemental Figure S1;[32]). Fluorescent staining of TSG-6 and hyaluronan (HA; an essential component of cartilage matrix and well-characterised TSG-6 ligand[22]) revealed 4 discrete populations of chondrocytes, corresponding to cells associated with TSG-6 staining alone, HA staining alone, staining for both molecules or lack of staining for either (Figure 1B). TSG-6 and HA were rarely found in association with the same cells (illustrated in Figure 1Bii). TSG-6 immunoreactivity was predominantly associated with chondrocytes (including cell clusters) in the most damaged regions of the tibial plateau (Figure 1 and Supplemental Figure S2). Use of an ‘object counting’ approach, to quantify fluorescent staining within individual zones of cartilage from three AMG patients (see Supplemental Table S2), confirmed the association of TSG-6 with cells in regions of significant cartilage loss (i.e. all zones within T1 and the superficial and middle zones of T2; Figure 1C and Supplemental Figure S2). Small percentages of TSG-6+ cells were also identified within macroscopically undamaged regions (UD1 and UD2) of all the donor cartilages analysed (Figure 1 and Supplemental Figure S2). Joint

**Figure 1.**
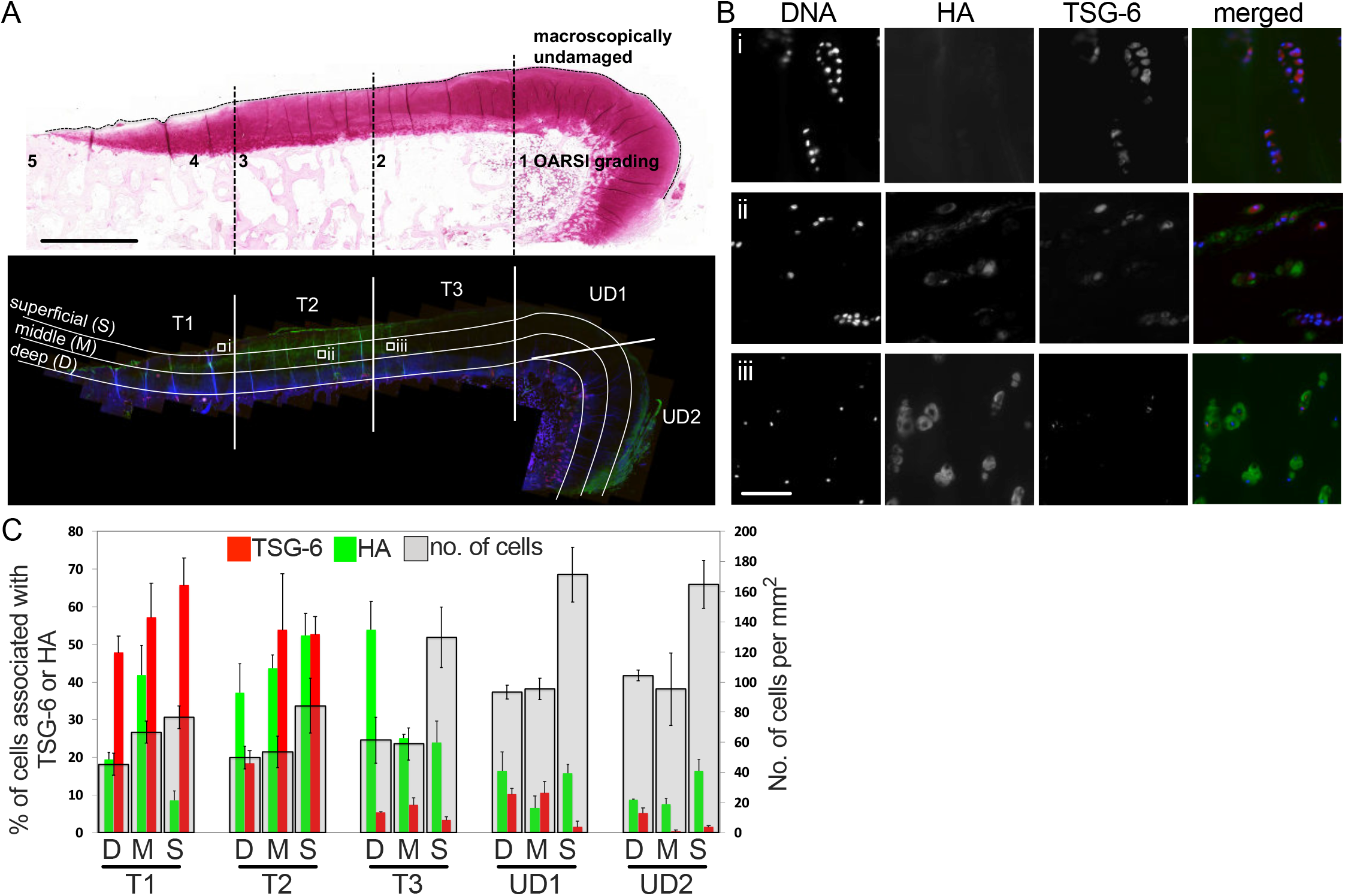
Differential localization of TSG-6 and HA in OA cartilage. **(A)** A representative section of medial tibial plateau (5 *μ*m thickness) from a patient with AMG stained with Safranin O (upper panel), DAPI (blue), a TSG-6-specfic antiserum RAH-1 (red) and HA-binding protein (green) (lower panel). Sections stained with Safranin O were graded according to the Osteoarthritis Research Society International (OARSI) scale. For quantitative analysis, full-thickness cartilage (OARSI grade ≤1.0) was denoted undamaged (UD) and was classified into UD1 (parallel to the bone/cartilage interface) and UD2 (perpendicular to the bone surface); cartilage between the UD region and the full thickness lesion (OARSI grade 5) was segregated into thirds (T1, T2 and T3) and all regions were further sub-divided longitudinally into superficial (S), middle (M) and deep (D) zones, where the latter was juxtaposed to the underlying subchondral bone (lower panel). Scale bar = 4 mm. **(B i-iii)** Representative images of AMG cartilage showing examples of cell populations associated with fluorescent staining of TSG-6 and/or HA; the regions from which these higher resolution images are captured are denoted i, ii and iii in the lower panel in **A**. Scale bar = 50 µm. **(C)** Semi-automated ‘object counting’ revealed the cell density and the percentage of cells associated with TSG-6 and HA staining within each of the zones defined in **A**. Data, which are from a single AMG donor and are representative of 3 distinct donors (see Supplementary Figure 2 for the other 2 donors), are presented as mean values from three sequential sections ± SEM.

tissue from patients with early-stage OA is not readily available, however, immunohistochemistry of rat knee cartilage 4-weeks after surgical induction of OA (ACLTpMMx) revealed cell- and matrix-associated TSG-6 staining both concomitant with cartilage fibrillation and in regions where lesions had not yet developed (Supplemental Figure 3). The expression of TSG-6 by chondrocytes in undamaged regions of human and rat OA cartilage led us to hypothesize that this protein may play a role in an attempted repair process.

### TSG-6 suppresses breakdown of human cartilage ***ex vivo***

In OA, cartilage damage is mediated by aggrecanase and collagenase enzymes, e.g. as a consequence of their elevated expression and/or reduced uptake by chondrocytes during inflammation[33-35]. Here we investigated whether TSG-6 can act directly on chondrocytes to counter the cytokine-mediated effects that drive cartilage breakdown. Cartilage explants from 59 OA patients undergoing TKA (Supplemental Table S2) were cultured with IL-1β/OSM in the absence or presence of either rhTSG-6 or the isolated Link module (Link_TSG6) and the extent of cartilage degradation determined by quantification of glycosaminoglycan (GAG) release. Due to its greater solubility and stability in solution, Link_TSG6 was tested at a wider range of concentrations than rhTSG-6 (≤33.3 µM and ≤10 µM, respectively, over 7 and 14 days). Both rhTSG-6 and Link_TSG6 dose-dependently inhibited cartilage breakdown, however, Link_TSG6 was more potent than the full-length protein (Figure 2A and Supplemental Figure 4A). For example, the effects of Link_TSG6 were statistically significant (p<0.001) at ≥0.1 *μ*M doses after 7 or 14 days treatment, while rhTSG-6 only had a significant effect after 7 days at the 1 *μ*M dose (p<0.05); at the highest dose of Link_TSG6 there was a substantial (∼70%) reduction in GAG loss.

**Figure 2.**
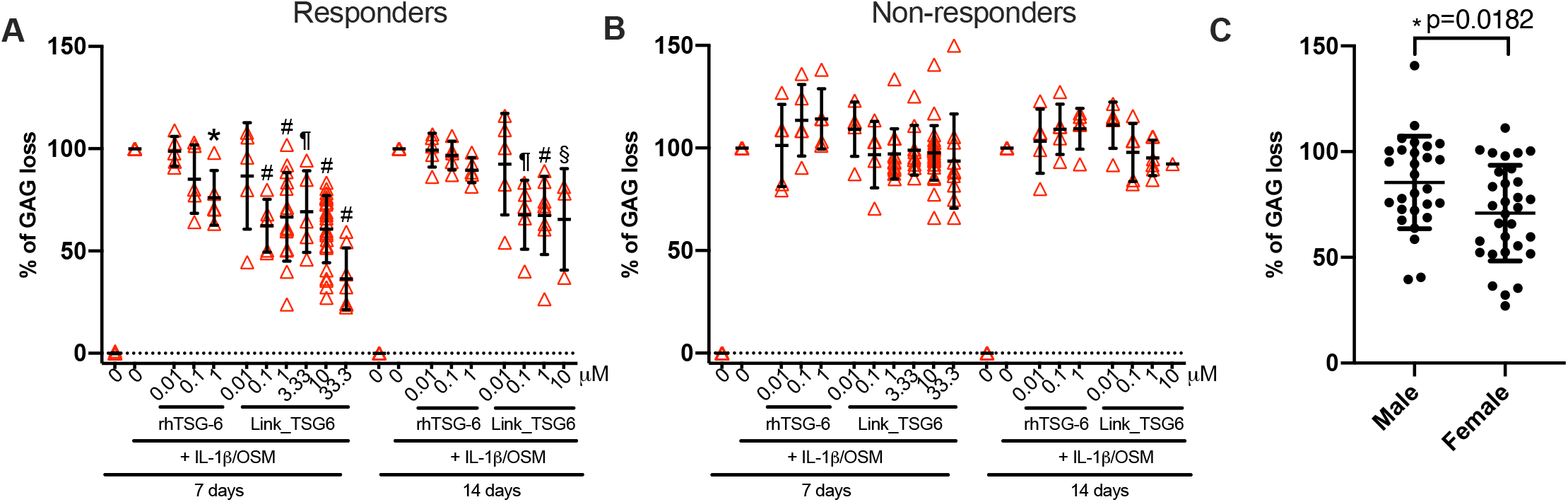
Link_TSG6 inhibits breakdown of OA cartilage *ex vivo*. Full thickness cartilage, dissected from the tibial plateaus of OA patients undergoing TKA, was incubated without or with 10 ng/ml IL-1β and 30 ng/ml Oncostatin M (OSM) and in the absence or presence of the indicated concentrations of rhTSG-6 or Link_TSG6; media were changed every seven days. Due to the limited solubility of rhTSG-6[43] the maximum concentration used was 1 µM, whilst Link_TSG6 was tested at up to 33.3 µM. Release of GAGs into the culture media after 7 or 14 days was determined using the Dimethylmethylene Blue assay and is expressed as a percentage of the GAG lost from each individual experimental sample upon treatment with cytokines alone (explant). Of the 59 donor cartilages tested, 30 were defined as ‘responders’ (i.e., a significant inhibitory effect over 7 days at ≤10 µM Link_TSG6) **(A)** and 29 were defined as ‘non responders’ to TSG-6 treatment **(B)**; 3-8 replicates per treatment per donor were analysed (see Supplementary Figure 4) and mean values for each donor are indicated by open triangles. Horizontal bars denote mean values for each treatment ±SEM. Data were analysed by one-way ANOVA with Tukey’s *post hoc* test \**p*<0.05, ^§^*p*<0.01, ^¶^*p*<0.001, ^#^*p*<0.0001 relative to the corresponding IL-1β/OSM alone control. **(C)** Percentage GAG loss of cartilages from female (n = 31) and male (n = 28) donors treated with 10 *μ*M Link_TSG6 in the presence of IL-1β/OSM; circles denote mean values for each donor and horizontal bars show the overall means ±SD. Data were analysed with a Mann-Whitney t-test; \**p*<0.05.

The inhibitory effect of Link_TSG6 varied greatly between donor cartilages. In this regard, thirty out of the 59 cartilages tested were responsive to Link_TSG6 treatment (Figure 2A; Supplemental Figure 4A), whereas in the other 29 there was no significant effect (Figure 2B; Supplemental Figure 4B). This suggests that cartilages (and their donors) can be stratified into Link_TSG6 ‘responder’ and ‘non-responder’ cohorts. Additionally, analysis of data from all cartilages treated with 10 *μ*M Link_TSG6 revealed a significantly greater reduction in cytokine-mediated matrix breakdown in tissues from female donors compared to those from male donors (Figure 2C). The finding that Link_TSG6 and rhTSG-6 inhibit cartilage breakdown *ex vivo* led us to further explore whether TSG-6 regulates catabolic pathways in human chondrocytes in OA cartilage and 3D pellet cultures.

### The expression of TSG-6 and MMP13 are negatively correlated in OA cartilage

To investigate the role of TSG-6 in OA cartilage we used RNAscope to quantitate endogenous *TSG-6* mRNA in individual chondrocytes and relate this to the expression of particular genes implicated in the OA disease process. Analysis of cartilage sections from 4 donors (Supplemental Table 2) revealed that *TSG-6* expression negatively correlates (p<0.0001) with *MMP13* expression at the single cell level; i.e. chondrocytes with high numbers of *TSG-6* transcripts typically have very few or no *MMP13* transcripts and *vice versa* (Figure 3A). This is indicative of distinct chondrocyte phenotypes (with different catabolic activities), where we have observed *TSG-6*^High^*MMP13*^Low^ and *TSG-6*^Low^*MMP13*^High^ cells in close proximity to each other within cartilage (Figure 3A, right panel). In contrast, data from the same 4 donors showed a positive correlation (p<0.0001) between *TSG-6* and *ADAMTS5* expression (Figure 3B). Together these results indicate that endogenous TSG-6 likely modulates the expression of catabolic enzymes in OA chondrocytes, potentially via the inhibition of inflammatory pathways.

**Figure 3.**
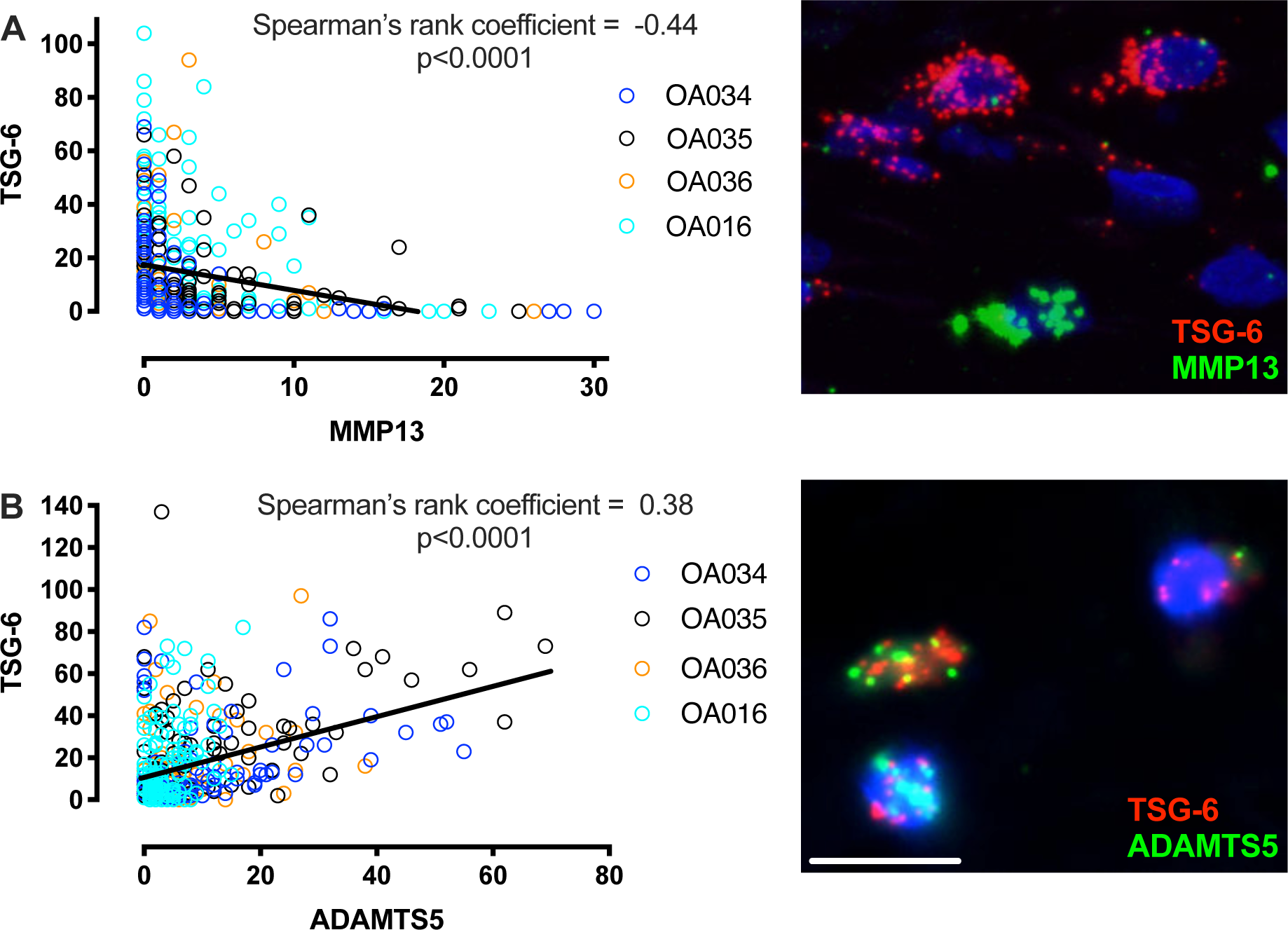
*TSG-6* mRNA expression in human OA cartilage and its correlations with *MMP13* and *ADAMTS5* expression. RNAscope was used to quantitate mRNA transcripts in individual chondrocytes within sections of damaged OA cartilage. Transcripts for **(A)** *TSG-6* and *MMP13* and **(B)** *TSG-6* and *ADAMTS5* were labeled using specific RNAscope probes in cartilages from 4 different donors; ≥ 100 cells per donor, expressing at least one transcript of either mRNA, were analysed. Left hand panels show the numbers of transcripts per cell for paired combinations of TSG-6 with **(A)** *MMP13* or **(B)** *ADAMTS5*; individual donor data are represented by coloured circles as indicated in the key. Spearman’s rank method was used to determine correlations and *p* values were calculated using Mann-Whitney U tests. Representative images of cells labeled with the indicated probe pairs are shown (right hand panels) with *TSG-6* transcripts in red and other transcripts in green; scale bar = 10 *μ*m. The significant negative correlation between *TSG-6* and *MMP13* expression levels (**A**; left hand panel) was also apparent when cartilages were analysed individually. There was a positive correlation between *TSG-6* and *ADAMTS5* expression levels (**B;** left hand panel) in only three out of four cartilages when analysed individually (data not shown).

### TSG-6 suppresses chondrocyte responses to pro-inflammatory cytokines

Given that rhTSG-6 and Link_TSG6 inhibited the breakdown of OA cartilage explants, we investigated the effects of these proteins on cytokine-induced expression of aggrecanase (ADAMTS4 and ADAMTS5) and collagenase (MMP13) enzymes in hMSC-derived chondrocytes in 3D pellet cultures. After 14-days in culture the hMSCs had differentiated into cells displaying a chondrocyte-like gene expression profile and had deposited a cartilage-like matrix with little or no TSG-6 expressed at the gene or protein level (Supplemental Figure S5). At this time point the chondrocytes were responsive to pro-inflammatory cytokines, where treatment for 24 h with 10 ng/ml IL-1α, IL-1β or TNF induced the expression of *ADAMTS4, ADAMTS5* and *MMP13* mRNAs (Figures 4 and 5). Thus, this pellet culture system represents a reasonable model of inflammatory pathways in OA cartilage in which to explore the activity of TSG-6. As shown in Figure 4,the inclusion of exogenous rhTSG-6 or Link_TSG6 protein suppressed IL-1β-induced expression of *ADAMTS4, ADAMTS5* and *MMP13* in a dose-dependent manner. Whereas 0.1 and 1.0 µM doses of Link_TSG6 reduced expression of *MMP13* to basal levels, the corresponding doses of rhTSG-6 were less effective. This demonstrates that the isolated Link module has more potent anti-catabolic/anti-inflammatory effects on chondrocytes than full-length TSG-6. This was supported by the observations that the 0.1 µM dose of Link_TSG6 was more effective than a 10-fold higher (1 *μ*M) dose of rhTSG-6 at inhibiting both IL-1α- and TNF-induced expression of *ADAMTS4, ADAMTS5* and *MMP13* (Figure 5).

**Figure 4.**
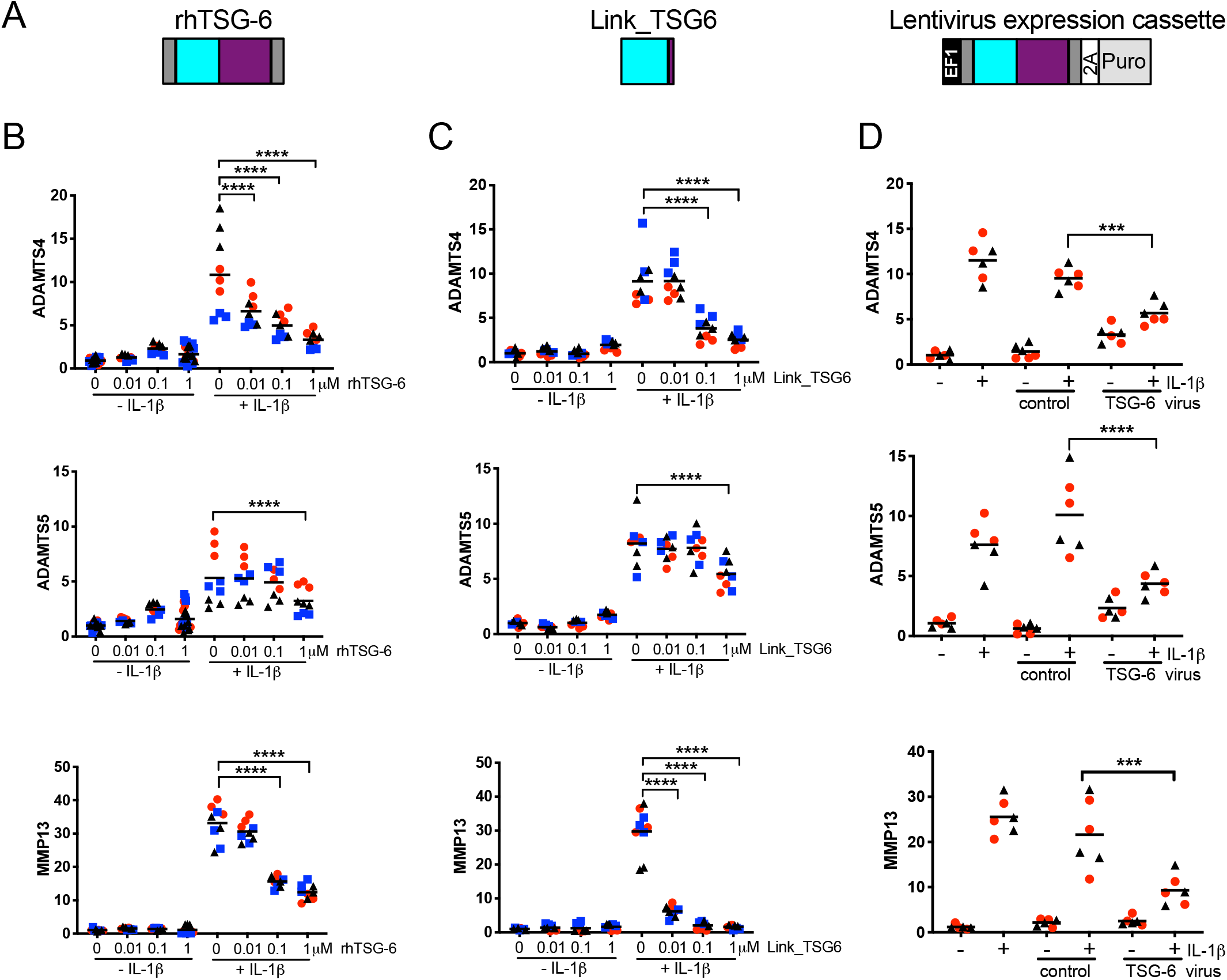
Full-length TSG-6 and Link_TSG6 suppress IL-1β-induced expression of *ADAMTS4, ADAMTS5* and *MMP13* in chondroctyes. **(A)** Schematic representations of the rhTSG-6 (left) and Link_TSG6 (centre) proteins that were added exogenously to chondrocyte pellet cultures and the expression cassette (right) within the pCDH-EF1-MSC-T2A-Puro vector (System Biosciences), which was used to generate lentivirus for endogenous over-expression of full-length TSG-6. hMSCs were cultured as 3D pellets in chondrogenic media for 14 days and then treated for 24 h, in the presence or absence of 10 ng/ml IL-1β with/without rhTSG-6 **(B)** or Link_TSG6 **(C)** at the concentrations indicated. hMSCs transduced with lentivirus **(D)** and constitutively expressing full-length TSG-6 (or transduced with control virus) were similarly cultured as chondrocyte pellets for 14 days and then incubated with or without IL-1β for 24 h. *ADAMTS4, ADAMTS5* and *MMP13* gene expression levels were determined by qPCR and are presented as fold-change relative to the no-addition control. Experiments were performed in triplicate with hMSCs from at least two donors (Donor A = red circles, Donor B = blue squares, Donor C = black triangles). Data from each experiment are shown (n=9-12 for rhTSG-6 and Link_TSG6 addition; n=6 for TSG-6 over-expression) with mean values indicated by horizontal lines. Data were analysed using two-way ANOVA with Tukey’s *post hoc* test; \*\*\**p*<0.001 and \*\*\*\**p*<0.0001 relative to the corresponding control with IL-1β.

**Figure 5.**
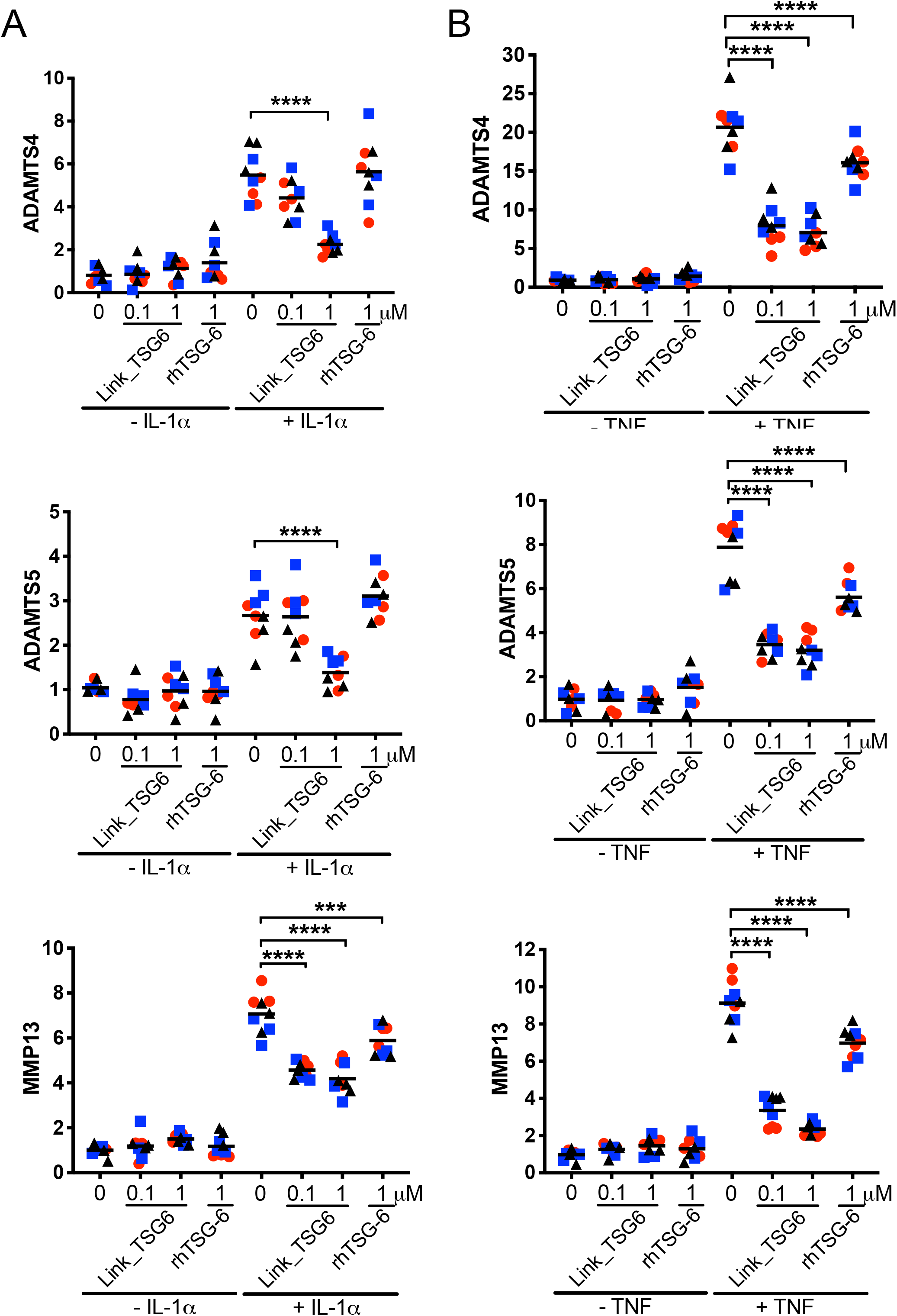
Link_TSG6 and rhTSG-6 suppress IL-1α- and TNF-induced expression of *ADAMTS4, ADAMTS5* and *MMP13* in chondroctyes. hMSCs were cultured as 3D pellets in chondrogenic media for 14 days and then treated for 24 h, in the presence or absence of 10 ng/ml IL-1α **(A)** or 10 ng/ml TNF **(B)** and with/without Link_TSG6 or rhTSG-6 at the concentrations indicated. *ADAMTS4, ADAMTS5* and *MMP13* gene expression levels were determined by qPCR and are presented as fold-change relative to the no-addition control. Experiments were performed in triplicate with hMSCs from three donors (Donor A = red circles, Donor B = blue squares, Donor C = black triangles). Data from each experiment are shown (n=9) with mean values indicated by horizontal lines. Data were analysed using two-way ANOVA with Tukey’s *post hoc* test; \*\*\**p*<0.001 and \*\*\*\**p*<0.0001 relative to the corresponding control with IL-1α or TNF.

In a complementary approach, endogenous full-length TSG-6 was over-expressed in hMSCs using lentivirus (Figure 4A). When these cells were differentiated into chondrocytes in 3D pellets their responses to IL-1β (Figure 4D), IL-1α and TNF (Supplemental Figure S6) were significantly reduced compared to controls. Collectively, this shows that expression of TSG-6 in human chondrocytes, or their treatment with TSG-6 proteins, effectively suppresses the expression of cartilage degrading proteases. Thus, we have identified a novel chondroprotective mechanism for TSG-6, via inhibition of inflammatory cytokine-mediated catabolic pathways, and shown that Link_TSG6 is considerably more potent than the full-length protein.

### Link_TSG6 reduces cartilage damage in the ACLTpMMx rat model of OA

Our *in vitro* and *ex vivo* data demonstrate that Link_TSG6 can potently suppress the cytokine-driven expression of proteolytic enzymes by chondrocytes and the loss of aggrecan from cartilage explants. We then evaluated the potential of Link_TSG6 to limit joint damage in OA using the rat ACLTpMMx model, where surgically induced joint destabilization gives rise to rapid cartilage degeneration[36,37]. Link_TSG6 (14, 40 and 120 µg doses) was administered via intra-articular (*i*.*a*.) injections at 7, 14 and 21 days post-surgery. At the 28-day endpoint histopathological scoring across seven parameters (Supplemental Table S3) revealed a dose-dependent reduction in damage to the medial tibia (Figure 6 and Supplemental Figure 7). The effects of Link_TSG6 on cartilage degeneration were particularly striking. At the highest dose (120 µg) there were significant reductions both in the severity and number of fibrillation/fissures in the cartilage (Figure 6A, D) and in the extent of cartilage erosion (Figure 6B, E). For example, in the latter, five vehicle-treated rats had a maximal erosion score of 4 (exposed bone) and one had a score of 0 (normal cartilage), whereas of the rats treated with 120 µg Link_TSG6 only one had a score of 4 and five had a score of 0. Furthermore, Link_TSG6 suppressed proteoglycan (PG) loss, where this effect was significant at the 14 *μ*g dose (Figure 6C, F). Overall these data indicate that intra-articular administration of Link_TSG6 has the potential to slow the OA disease process.

**Figure 6.**
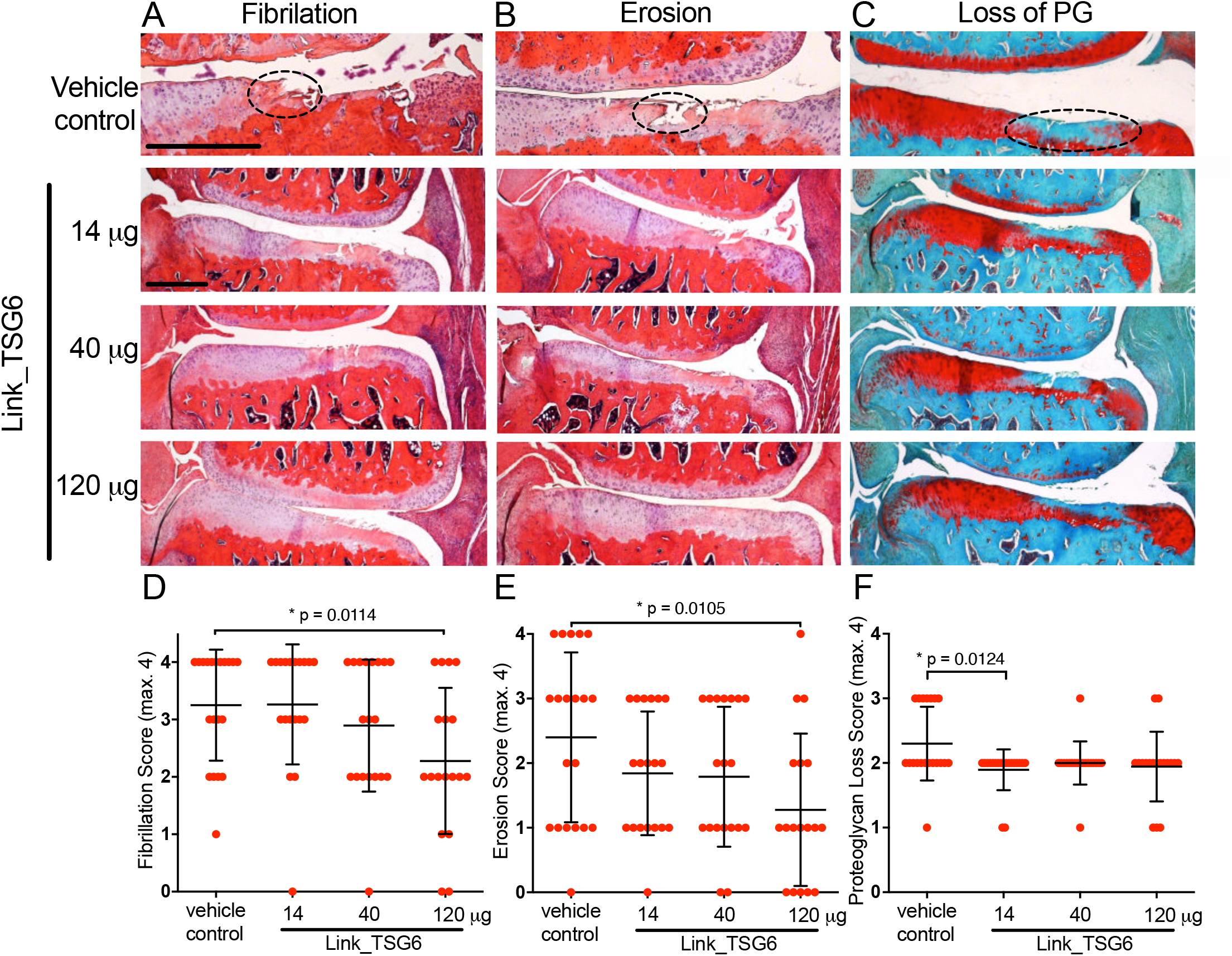
Link_TSG6 reduces cartilage damage *in vivo* in the ACLTpMMx rat. OA was induced in the right knees of male Lewis rats by transection of the anterior cruciate ligament (ACLT) and a partial (30%) medial meniscectomy (pMMx). Link_TSG6 treatment (or vehicle) was administered *i*.*a*. at 7, 14 and 21 days post-surgery, at the doses indicated. At the 28-day end point, joint tissues were fixed, decalcified and 7 µm sections were stained with Safranin O/Fast Green or Alcian Blue (pH 2.5), prior to blinded histopathological scoring by two individuals. Representative stained sections for each of the treatment groups are shown to illustrate **(A)** cartilage fibrillation, **(B)** erosion and **(C)** loss of proteoglycan (PG); pathological features are indicated with dashed ovals. The outcomes of scoring these parameters (on scales of 0 to 4; see Supplementary Table 3) for the medial tibiae are shown in **(D), (E)** and **(F)**, respectively. Data are shown as scatter plots for each treatment group (n=18-20), with the mean scores ± SD indicated by horizontal bars. Data were analysed by using t tests (Mann Whitney), where *p* values <0.05 are shown.

### Link_TSG6 suppresses tactile allodynia in the ACLTpMMx model

We investigated whether the reduced joint damage in Link_TSG6-treated animals described above (Figure 6) was associated with effects on a symptom of OA. ACLTpMMx rats were treated with Link_TSG6 (40 µg/week, *i*.*a*.) and, at the 4-week time point, touch-evoked pain (tactile allodynia) was assessed in the paws of surgically destabilized (right) and un-operated control (left) legs as an indicator of secondary hyperalgesia. Paw withdrawal responses for the un-operated legs (with or without Link_TSG6 treatment) were essentially identical to those of sham operated control animals (Figure 7A). However, in ACLTpMMx-operated legs (vehicle treated) there was a significant increase in sensitivity to punctate mechanical stimulus. This response was significantly reduced by intra-articular injection of Link_TSG6 (Figure 7B). Macroscopic evaluation of the medial tibiae (using digital segmentation to quantify cartilage damage, exposed bone and osteophyte/chondrophyte formation) at the 28-day study endpoint revealed extensive structural damage associated with ACLTpMMx surgery (Figure 7C,D). In Link_TSG6-treated animals the mean total lesion area was reduced by ∼30% (Figure 7D), which is consistent with the histological analysis of cartilage fibrillation and erosion in the dose-response study (Figure 6). The above *in vivo* data support the potential of intra-articular Link_TSG6 as a protein biological treatment for OA.

**Figure 7.**
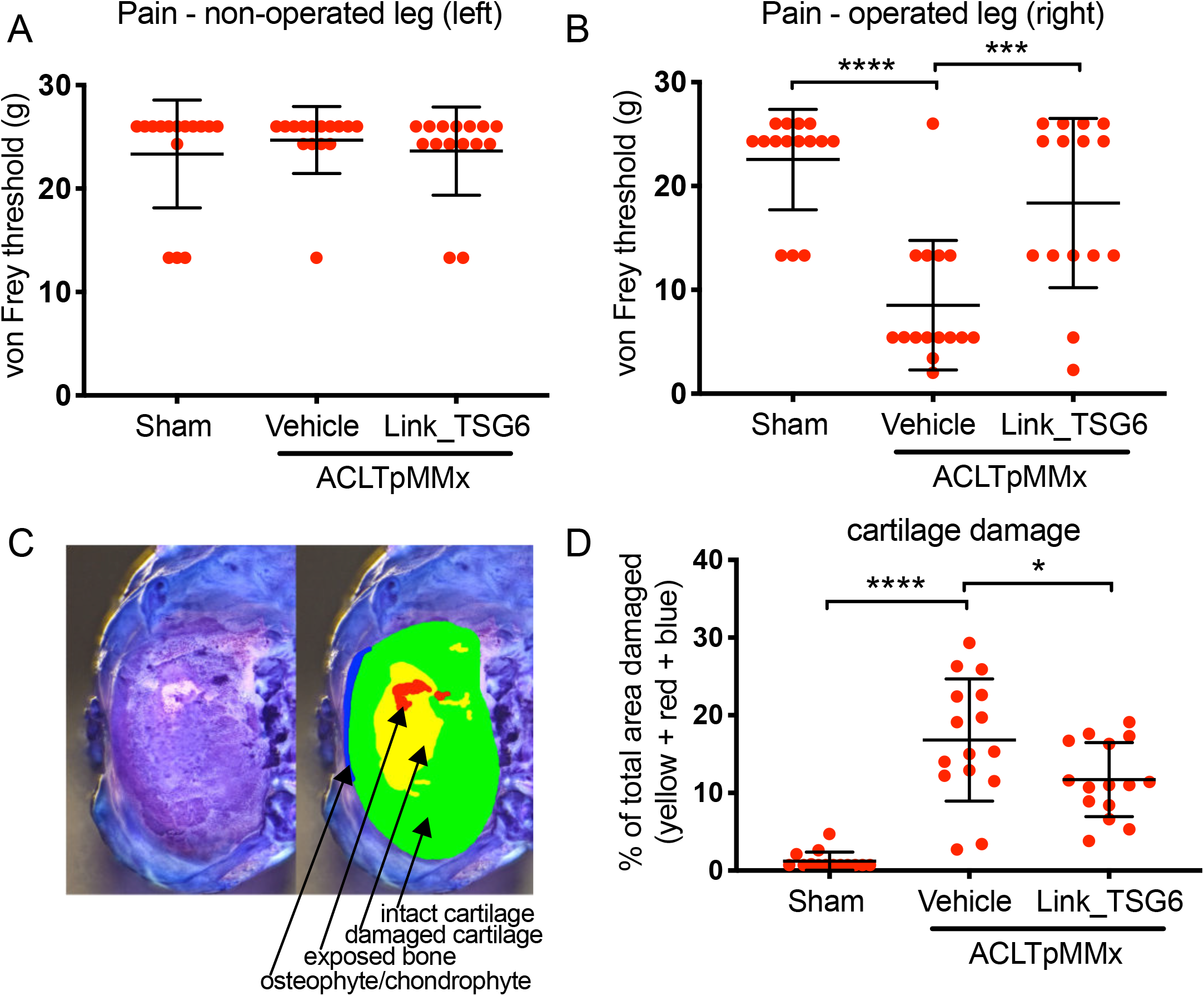
Link_TSG6 reduces tactile allodynia and cartilage lesion size in the ACLTpMMx rat. OA was induced in the right knees of male Lewis rats by ACLT and pMMx (30%). Link_TSG6 treatment (40 *μ*g) or vehicle was administered *i*.*a*. at 7, 14 and 21 days post-surgery. After 28 days tactile allodynia was assessed by determining withdrawal thresholds to von Frey hairs applied to hind paws of the **(A)** un-operated control legs and **(B)** operated legs. **(C)** Animals were then sacrificed, articular joint surfaces were stained with India Ink (left panel) and digital macroscopy (right panel) was used to determine the areas of intact cartilage (green), damaged cartilage (yellow), exposed bone (red) and osteophyte/chondrophyte formation (blue). (**D**) The total damaged area was determined (yellow + red + blue) as a percentage of total tibial plateau area, for each animal. Data in **(A), (B)** and **(D)** are presented as scatter plots for each treatment group (n=15), with mean values ± SD indicated by horizontal bars. Data were analysed by one-way ANOVA with Tukey’s *post hoc* test; \**p*<0.05, \*\*\**p*<0.001, \*\*\*\**p*<0.0001, relative to vehicle control.

## DISCUSSION

Here we have shown that TSG-6 acts through a novel chondroprotective mechanism, suppressing

inflammatory cytokine-mediated expression of the major aggrecanase and collagenase enzymes implicated in OA pathology. Thus, our observation that TSG-6 protein is largely associated with the most damaged regions of articular cartilage is consistent with it having an intrinsic function in tissue protection rather than driving the OA disease process[30,38]. Moreover, this aligns with a wealth of data on the reparative and anti-inflammatory effects of TSG-6 in a diverse range of animal models[22]. The role of inflammatory cytokines in OA pathogenesis has been an area of debate[39-41] but the recent clinical trial showing that Canakinumab reduces rates of knee and hip replacement[42] provides strong evidence that inhibition of IL-1β (and likely other cytokines) is a good therapeutic target.

The full-length TSG-6 protein is not suitable as a biological drug due to its poor solubility and the fact that it aggregates even at low concentrations[43]. On the other hand, not only can Link_TSG6 be used at much higher doses, it is also considerably more potent than rhTSG-6 at inhibiting the cytokine-induced breakdown of cartilage explants and suppressing the induction of ADAMTS4, ADAMTS5 and MMP13 expression in chondrocytes. This, along with the efficacy of intra-articular Link_TSG6 in a rat model of surgically-induced osteoarthritis, underpins its potential as a DMOAD. Our *in vivo* studies revealed that, in addition to its protective effects on cartilage, Link_TSG6 also reduces touch-evoked pain in the ACLTpMMx rat. At present the mechanism underlying the latter is unknown, but might involve the interaction of Link_TSG6 with chemokines[24,44-46]; further work is needed to explore this. Interestingly, we found no correlation between Link_TSG6’s effects on joint structure and tactile allodynia (data not shown), which is consistent with observations in human OA[47].

Our finding that ∼50% of human donor cartilages are responsive to Link_TSG6 (*ex vivo*) suggests it may have therapeutic utility for a large number of OA patients. Cartilage explants from female donors exhibited a greater treatment response compared to those from males, consistent with substantial evidence for sex differences in drug pharmacokinetics including in other arthritides[48,49]; clinical parameters (pain and function scores) did not correlate with the sex-associated difference in response to Link_TSG6 (data not shown). *A priori* determination of Link_TSG6 responder status (which may be influenced by factors such as chondrocyte phenotype and/or the extent and nature of cartilage damage) will be important in the context of future clinical trials. In this regard, it has been shown previously that stratification based on response to treatment is feasible in OA[50].

Our analysis of endogenous TSG-6 in OA cartilage (at both protein and RNA levels) indicates that it is a marker of chondrocyte phenotype, being expressed by a subset of cells. Importantly, *TSG-6* expression in the most damaged regions of cartilage and its inverse correlation with *MMP13* mRNA, suggest that it is associated with chondrocytes that have low catabolic activity against type II collagen. It seems likely that TSG-6 actively suppresses collagenase production in this context based on the 3D pellet culture data showing that rhTSG-6, or lentiviral overexpression of TSG-6, counter IL-1-/TNF-induced MMP13 expression. Currently, the mechanism by which TSG-6 inhibits cytokine-mediated pathways in OA chondrocytes is not understood. While there is no evidence that TSG-6 has a dedicated cell surface receptor, some of its anti-inflammatory activities have been found to be dependent on CD44 and may involve HA[22]; i.e. given that TSG-6-mediated HA crosslinking enhances its interaction with HA receptors[51-53]. However, in cartilage this mechanism of action seems unlikely since Link_TSG6 (which is more potent than rhTSG-6) only poorly enhances HA-CD44 interactions compared to the full-length protein[51] and our observation here that TSG-6^+^ chondrocytes have little or no HA staining.

Single cell analysis in OA cartilage revealed a positive correlation between *ADAMTS5* and *TSG-6* mRNA levels; here, our data indicate substantial variability in *ADAMTS5* expression at the cellular level. ADAMTS5 plays a role in cartilage homeostasis as well as in OA pathology[54,55]; thus, it is possible that TSG-6 regulates the expression of this aggrecanse and matrix turnover as a part of an attempted repair process. We do not know either the inflammatory or mechanical environment of the chondrocytes in the ‘endstage’ OA tissues used here. However, our pellet culture and explant studies indicate that TSG-6 inhibits aggrecanse production and activity in experimental settings with high levels of inflammatory cytokines. This is consistent with a recent study showing that deletion of the *Tsg6* gene in mice is associated with the rapid upregulation of proinflammatory and catabolic gene expression (*Adamts5, Ccl2* and *Il1a*) following joint destabilisation[38]. Moreover, in human cartilage the level of ADAMTS5 protein is, in part, dictated by its residence time in the tissue, which is dependent on LRP-1-mediated endocytosis by chondrocytes[34]; in OA, uptake of ADAMTS5 is impaired via cytokine-dependent shedding of LRP-1 leading to increased aggrecanase activity[35].

In summary, this study has identified a potential new treatment for osteoarthritis based on the Link module from human TSG-6. Link_TSG6 mimics the intrinsic anti-inflammatory and chondroprotective properties of the endogenous TSG-6 protein, as well as having greater potency, making it an attractive and novel target as a DMOAD.

## Data Availability

All data relevant to the study are included in the article or uploaded as supplementary information.

## Author Contributorship

Substantial contributions to the conception or design of the work (SPD, EB, AJP, SA, LCB, TL, MH, CMM, AJD), or the acquisition (SPD, NK, JLS, DPD, JMT, EB), analysis or interpretation of data (SPD, NK, JLS, EB, CMM, AJD).

Drafting the work or revising it critically for important intellectual content (all authors). Final approval of the version published (all authors).

Agreement to be accountable for all aspects of the work in ensuring that questions related to the accuracy or integrity of any part of the work are appropriately investigated and resolved (all authors.

## Funding and Acknowledgements

We acknowledge funding from Versus Arthritis (grants 18472, 20895, 21946 and 22277), the Medical Research Council (MRC) Confidence in Concept scheme and a Wellcome Trust Institutional Translational Partnership award. The Wellcome Trust Centre for Cell-Matrix Research is supported by core funding from the Wellcome Trust (203128/Z/16/Z). The Bioimaging Facility microscopes used in this study were purchased with grants from BBSRC, Wellcome Trust and the University of Manchester Strategic Fund; special thanks go to Roger Meadows and Steve Marsden for assistance and training. We also thank Pete Walker and Grace Bako (Histology Facility, FBMH, University of Manchester) for help with sample processing and sectioning. We thank Giles Hassall and Viranga Tilakaratna for the production of Link_TSG6 and rhTSG-6 proteins and Thomas Bissinger, Kerstin Stephan and Inge Berg-Scholl for expert technical assistance. Oncostatin-M (OSM) and IL-1α were generous gifts from Andrew Rowan (Newcastle).

Parts of this work have been presented at the following conferences and meetings: *Gordon Research Conference on Cartilage Biology & Pathology*, Les Diablerets, Switzerland, 2013; *World Congress on Osteoporosis, Osteoarthritis and Musculoskeletal Diseases*, Milan, Italy, 2015 (Kanakis *et al*. (2015) Preclinical evaluation of the Link module from the human TSG-6 protein: a potential new therapeutic for bone loss and cartilage damage in musculoskeletal disorders. *Osteoporosis International* **26**; S360); *OARSI 2016 World Congress*, Amsterdam, The Netherlands, 2016 (Day *et al*. (2016) A novel chondroprotective property of TSG-6 has therapeutic potential for OA. *Osteoarthr Cartil* **24**; S19-S20); *BSMB Spring Meeting*, Chester, UK, 2016 (Milner *et al*. (2016) TSG-6 protects cartilage and bone by modulating the activities of chondrocytes and osteoclasts: a potential therapeutic for musculoskeletal disorders. *Int J Exp Pathol* **97**; A26-A27); *Gordon Research Conference on Proteoglycans*, Andover, USA, 2016; *International Cartilage Repair Society World Congress*, Sorrento, Italy, 2016; *11*^*th*^ *International Conference on Hyaluronan*, Cleveland, Ohio, USA, 201*7; Gordon Research Conference on Proteoglycans*, Andover, USA, 2018; *Matrix Biology Europe 2018*, Manchester, UK, 2018; *FEBS – Extracellular Matrix: Cell Regulation, Epigenetics & Modeling*, Patras, Greece, 2018.

