## Supplementary material for "The recombinant Link module of human TSG-6 suppresses cartilage damage in models of osteoarthritis: a potential disease-modifying OA drug": Supplemnetal Material

[1]Wellcome Trust Centre for Cell-Matrix Research, [2]Faculty of Biology Medicine & Health, University of Manchester, Manchester Academic Health Science Centre, Manchester, UK; [3]Sanofi Aventis Deutschland GmbH, D-65926 Frankfurt, Germany; [4]Nuffield Department of Orthopaedics, Rheumatology & Musculoskeletal Sciences, University of Oxford, Oxford, UK; [5]Department of Orthopaedics, Stepping Hill Hospital, Stockport, UK; [6]Manchester Orthopaedic Centre, Manchester University Hospitals Foundation Trust, Manchester, UK.

***Correspondence to:***

Professor Anthony J Day, Wellcome Trust Centre for Cell-Matrix Research, Faculty of Biology Medicine & Health, University of Manchester, Michael Smith Building, Oxford Road, Manchester M13 9PT, UK;

Dr Caroline M Milner, Faculty of Biology Medicine & Health, University of Manchester, Michael Smith Building, Oxford Road, Manchester M13 9PT, UK;

#### Methods

##### Recombinant proteins

Full-length recombinant human TSG-6 (rhTSG-6) and Link_TSG6 were expressed in *Drosophila* S2 cells and *E. coli*, respectively, as described previously [1-3]. Oncostatin-M (OSM) and IL-1$\alpha$ were generous gifts from Andrew Rowan (Newcastle); all other cytokines were purchased from PeproTech.

##### Human tissue

Human tissues (Supplemental Table 2) were used with UK NHS Research Ethics Committee approval and with subjects' informed consent. Medial tibial plateaus were obtained from patients with anteromedial gonarthrosis (AMG) undergoing unicompartmental knee arthroplasty (UKA; Nuffield Orthopaedic Centre, Oxford; NHS REC 09/H0606/11); whole tibial plateaus were obtained from patients with symptomatic OA undergoing total knee arthroplasty (TKA; Stepping Hill Hosptital, Stockport, or Manchester University Hospitals Foundation Trust, Manchester; NHS REC 13/EM/0388). Anonymised donor identifier codes (known only to the research team) are used throughout.

##### Histological and fluorescent staining of human OA cartilage

Medial tibial plateaus from AMG patients were grossly cut into full-length segments (posterior to anterior; encompassing both macroscopically normal and damaged regions of cartilage), decalcified with 10% (v/v) EDTA or 5% (v/v) formic acid and embedded in paraffin wax. Some sections (5 μm) of cartilage were stained with hemotoxylin and eosin (H&E) or 1% (w/v) Alcian Blue (pH 1.0 or pH 2.5; to stain highly and lowly sulfated glycosaminoglycans, respectively).

For each donor, ≥3 sequential sections (5 $\mu$m) were cut through the full length of these segments and processed for fluorescence microscopy; an adjacent section was stained with 0.1% (w/v) Safranin O (to stain proteoglycans) and graded according to the Osteoarthritis Research Society International (OARSI) scale. Full-thickness cartilage (OARSI grade ≤1.0) was denoted undamaged (UD) and was sub-divided into UD1 (parallel to the bone/cartilage interface) and UD2 (perpendicular to the bone surface); cartilage between the UD region and the full thickness lesion (OARSI grade 5) was segregated into thirds (T1, T2 and T3) and all regions were further sub-divided longitudinally into superficial (S), middle (M) and deep (D) zones, where the latter was juxtaposed to the underlying subchondral bone (see Figure 1A).

For fluorescent staining, TSG-6 was detected using RAH-1 antiserum [4] (diluted 1:1000 in PBS containing 2% (v/v) FBS) followed by Alexa Fluor 555-conjugated goat anti-rabbit IgG (1:500; Life Technologies), hyaluronan (HA) was detected with biotinylated HA-binding protein (bHABP, 250 $\mu$g/ml; Seikagaku [5]) followed by Alexa Fluor 488-conjugated streptavidin (1:500; Life Technologies) and DNA was stained with DAPI. Sections were mounted on glass with ProLong Gold (Thermo Fisher Scientific). Images were collected on an Olympus BX51 upright microscope using 4x/0.13 or 10x/0.30 UPlan FLN objectives and captured with a Coolsnap ES camera (Photometrics) using MetaVue Software (v7.8; Molecular Devices). The Stitching plugin (Fiji, ImageJ [6]) was used to reconstruct a single, high resolution fluorescent image (in three channels) spanning the full length of each knee tissue section; alternatively complete sections were scanned and automatically stitched together using a Panoramic 250 Flash digital slide scanner (3DHISTECH). Within these images, each zone (T1_S, T1_M, T1_D etc.) was split into greyscale representations of of the three component fluorescent channels and quantitative analysis was carried out based on semi-automated 'object counting' using the 3D Objects Counter plugin (Fiji, ImageJ[7]). Total cell densities (cells/mm$^2$) were determined by counting DAPI-stained nuclei (blue channel); cells associated with TSG-6 and/or HA staining were identified in the red and/or green channel, respectively.

**Human cartilage explant assay**

Full-thickness, macroscopically undamaged, cartilage was dissected from whole tibial plateaus within 2 h of TKA, cut into approx. 2 x 2 x 2 mm cubes and incubated in serum-free DMEM (with penicillin, streptomycin and nystatin) in 5% $CO_2$, 95% air at 37°C. After 24 h, explants were transferred to 24-well plates (3 cubes/well) and incubated in the absence or presence of 10 ng/ml IL-1$\beta$ and 30 ng/ml OSM ± rhTSG-6 or Link_TSG6 at the concentrations indicated for up to 21 days; media were changed every 7 days. Glycosaminoglycan (GAG) release into the media, as an indicator of cartilage degradation, was quantified using the Dimethylmethylene Blue (DMB) assay [8]; the residual GAG content of cartilage tissue was determined using the same assay following overnight digestion with papain (Sigma Aldrich) at 65ºC.

**RNAscope analysis of human OA cartilage**

Damaged cartilage from the tibial plateaus of OA patients (T1 and T2 regions only; see Figure 1) was manually dissected from the underlying bone, fixed in 10% (v/v) formalin, then embedded in paraffin wax. Sections (5 $\mu$m) were processed for RNAscope according to the manufacturer’s protocol (RNAscope® Multiplex Fluorescent v2 Reagent Kit, Advanced Cell

Diagnostics) with the following probe sets: Hs-TNFAIP6-C1 (for TSG-6), Hs-MMP13-C3 (for MMP13) and Hs-ADAMTS5-C2 (for ADAMTS5); tissue compatibility was confirmed in each case. Images were acquired on an Olympus IX83 inverted microscope using UV, Green-Yellow and Red Lumencor LED excitations, a 60x 1.42 Plan Apo objective and the Sedat Quad 89000 filter set (Chroma Technology). Images were collected using an R6 CCD camera (Qimaging) with a Z optical spacing of 0.2 μm and Metamorph v7.8.4.0 software (Molecular Devices); maximum intensity projections were generated using ImageJ [9].

**Preparation and characterization of chondrocyte 3D pellet cultures**

All cell culture was carried out in 5% $CO_2$, 95% air at 37°C. Human mesenchymal stem/stromal cells (hMSC) were isolated from bone marrow mononuclear cell aspirates of donors aged 19-30 years (StemCell Technologies) by adherence to plastic over 24 h. Cells were expanded as monolayers in either Human MesenCult Proliferation Medium (StemCell Technologies) or Mesenchymal Stem Cell Growth BulletKit Medium (Lonza), containing 5 ng/ml FGF-2 (PeproTech). Cells were passaged from the point when they reached confluence (passage 1; P1) and differentiation to chondrocytes was initiated at P4 [10,11]. Briefly, hMSCs ($5 \times 10^5$ cells in 1 ml chondrogenic medium; high glucose DMEM containing 100 μg/ml sodium pyruvate (Sigma), 100 ng/ml TGFβ3 (PeproTech), 100 nM dexamethasone, 1x ITS+1, 40 μg/ml proline and 25 μg/ml ascorbate-2-phoshapte (all from Sigma-Aldrich)) were pelleted in 15 ml conical tubes (250 x *g*, 5 min). Pellets were maintained in chondrogenic medium, with media changed every 2 days, for up to 28 days.

For gene expression analysis pellets were disrupted (using a mortar and pestle and Molecular Grinding Resin (G-Biosciences)) in RNeasy Lysis Buffer, and lysates were homogenized using a QIAshredder column and RNA was purified according to the manufacturer's instructions (QIAGEN). cDNA was synthesized from 1 μg RNA using a High Capacity cDNA Reverse Transcription Kit (Life Technologies). Quantitative PCR (qPCR) was performed with the primer pairs shown in Supplemental Table 1 using PrecisionPLUS Master Mix with SYBR green (Primerdesign); fold changes in gene expression were determined relative to corresponding control conditions or relative to GAPDH using the $2^{-\Delta\Delta Ct}$ or $2^{-\Delta Ct}$ methods, respectively. Expression of *SOX9* (encoding a transcriptional activator of chondrocyte differentiation and promoter of chondrocyte viability), *COL2A1* and *ACAN* (encoding collagen II and aggrecan, respectively; major structural components of cartilage matrix), *COL1A1* (encoding collagen I, which has low abundance in mature cartilage) and *TSG-6* were determined.

For analysis of matrix, 14-day pellets were fixed, embedded in paraffin wax and processed for histological staining or fluorescence microscopy. Sections of pellets (5 μm) were stained with 0.1% (w/v) Safranin O or with 1% (w/v) Alcian Blue (pH 1.0 or pH 2.5). Brightfield images were collected on an Olympus BX63 upright microscope using a 4x or 10x 0.75 UApo/340 objective and captured and white-balanced using a DP80 camera (Olympus) in colour mode with CellSens Dimension v1.16 (Olympus). Images were then processed and analysed using Fiji ImageJ (http://imagej.net/Fiji/Downloads). In fluorescence analyses, chondrocytes were localized by staining nuclei with DAPI; HA was detected with biotinylated-HA-binding protein (bHABP)/Alexa Fluor 488-streptavidin and TSG-6 with RAH-1 antiserum/Alexa Fluor 555-goat anti-rabbit IgG. Fluorescent images were collected on an Olympus BX51 upright microscope using 4x/0.13 or 10x/0.30 UPlan FLN objectives and captured with a Coolsnap ES camera (Photometrics) using MetaVue Software (v7.8; Molecular Devices).

**Overexpression of TSG-6 in hMSC-derived chondrocytes using lentivirus**

The cDNA sequence encoding human TSG-6 was cloned into the *Xba*I and *Eco*RI sites of the pCDH-EF1-MSC-T2A-Puro vector (System Biosciences) to generate a transfer vector. Lentiviral particles were produced by co-transfection of HEK 293T cells with the TSG-6 transfer vector (or empty vector as control), the packaging plasmid psPAX2 and the envelope plasmid pMD2.G (Addgene), according to the supplier's protocol. hMSCs were infected with lentivirus at P2, subjected to selection with puromycin (3 μg/ml) for 3 days and then differentiated into chondrocytes in 3D pellet culture for 14 days, prior to incubation with cytokines for 24 h.

**Analysis of aggrecanase and collagenase gene expression in chondrocyte pellet cultures**

Chondrocyte 3D pellet cultures were generated from human MSCs (hMSC) and characterized as described above. Fourteen-day pellet cultures (maintained in chondrogenic medium, with media changed every 2 days) were treated with/without 10 ng/ml cytokine (IL-1α, IL-1β or TNF) in the absence/presence of rhTSG-6 or Link_TSG6 for 24 h. mRNA was extracted and qPCR was performed using the primer pairs listed in Supplemental Table 1 for *ADAMTS4*, *ADAMTS5*, *MMP13*; fold changes in gene expression were determined relative to the corresponding 'no cytokine' control using the $2^{-\Delta\Delta Ct}$ method. Fourteen-day 3D pellets generated from hMSCs transduced with lentivirus, to mediate overexpression of TSG-6 (see above), were also treated with cytokines and analysed by qPCR as above.

**ACLTpMMx rat model of OA**

All procedures involving experimentation on animals were performed according to Directive 63/2010 of the European Commission implemented into German Animal Welfare Legislation and are registered by the Competent Authority (District Government in Darmstadt) under No. HMR-3/ Anz. 37. Rats were housed in solid bottom polycarbonate type IVS cages, with hard wood bedding and wood wool as enrichment, one animal per cage and with access to food and water *ad libitum*. Animals were randomly allocated to experimental groups. Prior to surgery, rats were anaesthetized with isofluran. Osteoarthritis (OA) was induced in 12-week old, male Lewis rats (Charles River) by anterior cruciate ligament transection (ACLT) with partial (30%) medial meniscectomy (pMMx) [12,13]. The right knee was surgically destabilized and the left knee served as sham-operated control; in the latter group the knee joint capsule was opened, the patella relocated and the incision closed. Treatments were administered by intra-articular (*i.a.*) injection (50 μl) at the doses and time points indicated; animals were euthanized (by carbon dioxide followed by exsanguination) at 28 days post-surgery for histological and macroscopic evaluations of the medial tibiae. Prior to euthanasia, tactile allodynia was assessed by withdrawal thresholds to von Frey filaments applied to the plantar surface of hind paws [14]. Fixed joints were decalcified and 7 μm sections of paraffin-embedded tissue stained with 0.1% (w/v) Safranin O/Fast Green or 1% (w/v) Alcian Blue, pH 2.5, prior to blinded histopathological scoring of joint damage by 2 independent observers using a scale from 0 to 4 for each parameter [15] (see Supplementary Table 3). Alternatively, joint surfaces were stained with India ink for blinded macroscopic analysis of pathology via digital quantification with the following segmentation parameters: cartilage damage area, exposed bone area and osteophyte/chondrophyte area. Immunohistochemical staining of TSG-6 protein in knee joint sections was conducted using RAH-1 antisera [4].

**Statistical analyses**

Data were analyed using one- or two-way ANOVA, with Tukey's or Dunn's *post hoc* multiple comparisons test. A t-test (Mann-Whitney) was used for comparison of individual pairs of data; $p$ values of <0.05 were considered significant. In the ACLTpMMx model, data for animals that were identified as outliers using the Grubbs' test were excluded from analyses. Statistical analyses were performed with GraphPad Prism 7 or StatsDirect software.

**Supplemental Tables**

**Supplemental Table 1: qPCR primers.**

| Gene | Sequence (5' – 3') | $^{\$}T_a$ |
|---|---|---|
| *TSG-6* | Forward - CATATGGCTTGAACGAGCAGC | 57 |
| | Reverse - CTTTGCGTGTGGGTTGTAGC | 57 |
| *ADAMTS4* | Forward - CATGTGCAACGTCAAGGCTC | 57 |
| | Reverse - CACCACCAAGCTGACAGGAT | 57 |
| *ADAMTS5* | Forward - GCCTCTCCCATGACGATTCC | 57 |
| | Reverse - CCAGGATCTGCTTTCGTGGT | 57 |
| *MMP13* | Forward - AGGAGCATGGCGACTTCTAC | 57 |
| | Reverse - CAAGACCTAAGGAGTGGCCG | 57 |
| #*GAPDH* | Forward - CTCCTGTTCGACAGTCAGCC | 57 |
| | Reverse - CCCAATACGACCAAATCCGTTG | 57 |
| *SOX9* | Forward - GCTGCTGGGAAACATTTGCACTCT | 57 |
| | Reverse - GGGCACTTATTGGCTGCTGAAACA | 57 |
| *ACAN* | Forward - TTCAGTGGCCTACCAAGTGGCATA | 57 |
| | Reverse - AGCCTGGGTTACAGATTCCACCAA | 57 |
| *COL1A1* | Forward - CAGCCGCTTCACCTACAGC | 60 |
| | Reverse - TTTTGTATTCAATCACTGTCTTGCC | 60 |
| *COL2A1* | Forward - GGCAATAGCAGGTTCACGTACA | 60 |
| | Reverse - CGATAACAGTCTTGCCCCACTT | 60 |
| ∞*GAPDH* | Forward - ATGGGGAAGGTGAAGGTC | 60 |
| | Reverse - TAAAAGCAGCCCTGGTGACC | 60 |

$^{\$}T_a$ = annealing temperature; for all reactions the amplification programme was 10 min at 95°C followed by 40 cycles of 95°C for 30 s, $T_a$ for 30 s and 72°C for 30 s. Amplicon disassociation constants were measured after each qPCR reaction· #Primers for GAPDH with $T_a$ = 57ºC· ∞Primers for GAPDH with $T_a$ = 60ºC

**Supplemental Table 2. Anonymised identifier codes, sexes and age ranges of cartilage donors**

| Donor Code | Sex | Age[∞] | Donor Code | Sex | Age[∞] |
|---|---|---|---|---|---|
| OA001* | M | 70-74 | OA034[¶] | F | 75-79 |
| OA002* | M | 70-74 | OA035[¶] | M | 70-74 |
| OA003* | F | 60-64 | OA036[¶] | F | 70-74 |
| OA004[#R] | F | 75-79 | OA037[#N] | M | 65-69 |
| OA005[#N] | M | 75-79 | OA038[#R] | F | 55-59 |
| OA006[#N] | M | 80-84 | OA039[#R] | F | 55-59 |
| OA007[#R] | F | 75-79 | OA040[#R] | F | 60-64 |
| OA008[#R] | M | 60-64 | OA041[#N] | M | 60-64 |
| OA009[#R] | F | 75-79 | OA042[#N] | M | 65-69 |
| OA010[#R] | M | 65-69 | OA043[#R] | F | 75-79 |
| OA011[#N] | F | 75-79 | OA044[#N] | M | 50-54 |
| OA012[#N] | M | 60-64 | OA045[#N] | M | 75-79 |
| OA013[#N] | M | 70-74 | OA046[#R] | F | 75-79 |
| OA014[#R] | F | 60-64 | OA047[#R] | F | 65-69 |
| OA015[#R] | M | 70-74 | OA048[#R] | F | 65-69 |
| OA016[¶] | F | 65-69 | OA049[#N] | F | 70-74 |
| OA017[#N] | M | 70-74 | OA050[#R] | F | 70-74 |
| OA018[#R] | M | 75-79 | OA051[#R] | M | 55-59 |
| OA019[#R] | F | 60-64 | OA052[#N] | M | 75-79 |
| OA020[#R] | M | 75-79 | OA053[#N] | F | 55-59 |
| OA021[#N] | F | 75-79 | OA054[#R] | M | 65-69 |
| OA022[#N] | F | 70-74 | OA055[#N] | M | 70-74 |
| OA023[#N] | F | 75-79 | OA056[#R] | M | 75-79 |
| OA024[#R] | M | 65-69 | OA057[#R] | F | 70-74 |
| OA025[#R] | M | 60-64 | OA058[#N] | F | 60-64 |
| OA026[#N] | M | 75-79 | OA059[#N] | F | 50-54 |
| OA027[#N] | M | 65-69 | OA060[#N] | F | 80-84 |
| OA028[#N] | F | 70-64 | OA061[#N] | F | 60-64 |
| OA029[#R] | M | 85-89 | OA062[#R] | F | 65-69 |
| OA030[#N] | M | 65-69 | OA063[#N] | M | 55-59 |
| OA031[#R] | F | 75-79 | OA064[#R] | F | 50-54 |
| OA032[#N] | M | 80-84 | OA065[#R] | F | 65-69 |
| OA033[#R] | M | 65-69 | OA066[#N] | M | 55-59 |

[∞]Age range of donor. Donor cartilages used in: *immunofluorescence; [¶]RNAscope; [#]explant assays: R = Responder or N = Non-responder (i.e., a significant or non-significant inhibitory effect, respectively, over 7 days at ≤10 μM Link_TSG6).

**Supplemental Table 3: Histopathological scoring of joint damage in the rat ACLTpMMx model of OA.**

| *Parameter* | *Observed histopathology* | [¶]*Score* |
|---|---|---|
| Chondrocyte changes in superficial cartilage | normal | 0 |
| | swelling of chondrocytes | 1 |
| | focal loss of chondrocytes | 2 |
| | loss of chondrocytes in some regions | 3 |
| | almost complete loss of chondrocytes | 4 |
| Fibrillation and/or fissures in cartilage | normal | 0 |
| | minor surface irregularities | 1 |
| | widespread fibrillation and/or 1 small fissure | 2 |
| | 2 small or 1 medium fissures | 3 |
| | 3 small, 2 medium, or 1 large fissures | 4 |
| Cartilage erosion | normal | 0 |
| | erosions of the superficial zone | 1 |
| | erosions extending into intermediate zone | 2 |
| | erosions extending into deep zone | 3 |
| | exposure of bone | 4 |
| Loss of proteoglycan | normal | 0 |
| | staining reduced in 1-25 % of joint area | 1 |
| | staining reduced in 26-50 % of joint area | 2 |
| | staining reduced in 51-90 % of joint area | 3 |
| | staining reduced in >90 % of joint area | 4 |
| Loss of chondrocytes in middle/deep cartilage | normal | 0 |
| | slight focal decrease | 1 |
| | moderate decrease in cells | 2 |
| | widespread decrease (≥50% cartilage area) | 3 |
| | almost complete loss of cells | 4 |
| Synovial changes including synovitis | normal | 0 |
| | increase in number of lining cells | 1 |
| | thickening of sub-synovial tissue | 2 |
| | infiltration of few inflammatory cells | 3 |
| | infiltration of many inflammatory cells | 4 |
| Sclerosis of subchondral bone | normal | 0 |
| | beginning increase in thickness | 1 |
| | moderate increase | 2 |
| | marked increase | 3 |
| | no marrow space left in > half of the area | 4 |

[¶]Maximal Total Sum Score = 28

#### Supplemental Figure Legends

**Supplemental Figure S1. Cartilage from the tibial plateau of patients with anteriomedial gonarthrosis (AMG) shows a characteristic pattern of erosion.** Medial tibial plateaus from patients with AMG undergoing unicompartmental knee arthroplasty were cut into segments from posterior to anterior through the region of maximum cartilage loss; tissue was fixed, decalcified and embedded in paraffin wax. Sequential 5 μm sections were cut, adhered to glass slides and subjected to **(A)** fluorescence staining of cell nuclei (DAPI; blue), HA (biotinylated-HABP/Alexa Fluor 488-streptavidin; green) and TSG-6 (RAH-1/Alexa Fluor 555-goat anti-rabbit IgG; red), **(B)** Hematoxylin and eosin staining, **(C)** Alcian Blue pH 2.5 staining of lowly sulphated glycoaminoglycans (GAGs) or **(D)** Alcian Blue pH 1.0 staining of highly sulphated GAGs. The locations of macroscopically normal cartilage (at the posterior), through progressively more damaged tissue to a full thickness lesion with exposed bone (towards the anterior) can be seen in the images shown, which are representative of 3 donors analysed. Scale bars = 4 mm.

**Supplemental Figure S2. Quantitation of TSG-6 and HA in full-length medial tibial plateau sections from patients with OA.** Representative sections of medial tibial plateau (5 μm thickness) from 2 patients with AMG (upper panels in **A** and **B**) were stained with DAPI (blue), TSG-6-specfic antiserum RAH-1 (red) and HA-binding protein (green). Sections stained with Safranin O were graded according to the Osteoarthritis Research Society International (OARSI) scale; not shown. For quantitative analysis, full-thickness cartilage (OARSI grade ≤ 1.0) was denoted undamaged (UD) and was sub-divided into UD1 and UD2; cartilage between the UD region and the full thickness lesion (OARSI grade 5) was segregated into thirds (T1, T2 and T3) and all regions were further sub-divided longitudinally into superficial (S), middle (M) and deep (D) zones, where the latter was juxtaposed to the underlying subchondral bone; scale bar = 4 mm. Lower panels of **A** and **B** show the results of semi-automated 'object counting' for the two patients where data are presented as mean values from three sequential sections ± SEM.

**Supplemental Figure S3. TSG-6 is expressed by chondrocytes in the ACLTpMMx rat model of OA.** Joint tissues were collected from rats at 4 weeks post ACLTpMMx surgery. Immunohistochemical staining was conducted with RAH-1 antisera [4] to detect TSG-6; (**A**) low- and **(B,C)** high-magnification images are shown. Chondrocyte- and matrix-associated TSG-6 immunoreactivity (examples indicated with arrows and asterisks, respectively) were detected **(B)** in the absence of cartilage lesions as well as **(C)** in fibrillated tissue**.** Images are representative of 20 rat joints analysed. All scale bars = 50 μm

**Supplemental Figure S4. Analysis of cartilage explants treated with TSG-6 proteins stratifies patients into responder and non-responder cohorts**. Cartilages from 59 OA patients (undergoing total knee arthroplasty) were incubated without or with 10 ng/ml IL-1β and 30 ng/ml oncostatin M (OSM) in the absence or presence of rhTSG-6 or Link_TSG6 (at the concentrations indicated) for 7 or 14 days. Cartilage breakdown was quantitated based on glycosaminoglycan (GAG) release measured using the Dimethylmethylene Blue (DMB) assay. Data are shown for explants from donors that are **(A)** responsive (n = 30) or **(B)** non-responsive (n = 29) to ≤10 μM Link_TSG6 treatment for 7 days; data are presented for individual experiments, colour-coded by donor.

**Supplemental Figure S5. Characterization of human chondrocyte pellet cultures.** Bone marrow-derived hMSCs were grown in monolayer, then transferred (at P4) to 3D pellets and cultured in chondrogenic media for up to 28 days. **(A-E)** mRNA was extracted from cells in monolayer (P2) and from 3D pellets at the indicated times. Gene expression of **(A)** *SOX9*, **(B)** *COL2A1*, **(C)** *ACAN*, **(D)** *COL1A1* and **(E)** *TSG-6* were determined by qPCR, using the primers in Supplemental Table 1. Expression levels relative to GAPDH were calculated using the $2^{-\Delta ct}$ method. Representative data from one donor (Donor A) are illustrated (presented as mean values (n=3) ± SEM), where all 4 donors showed similar gene expression profiles, e.g. with elevated expression of *SOX9*, *ACAN* and *COL2A1* and low expression of *COL1A1* at 14 days. At the 14-day time point the cells had a chondrocyte-like gene expression profile and expressed low levels of endogenous TSG-6. **(F)** 14-day pellets were fixed and processed for histological staining or fluorescence microscopy. Sections of chondrocyte pellets (5 μm) were stained with Safranin O or with Alcian Blue (pH 1.0 or pH 2.5) to detect proteoglycans and highly or lowly sulphated glycosaminoglycans, respectively (insets show sections of entire pellets). The localizations of chondrocytes (nuclear DNA stained with DAPI), HA (detected with biotinylated HABP/Alexa Fluor 488-streptavidin) and TSG-6 (detected with RAH-1/Alexa Fluor 555-goat anti-rabbit IgG) were determined by immunofluorescence microscopy; scale bar = 100 μm. These data demonstrate that 14-day pellet cultures produce a matrix rich in proteoglycans and glycosaminoglycans, with little or no endogenous TSG-6 protein.

**Supplemental Figure S6. TSG-6 overexpression suppresses IL-1α- and TNF-induction of catabolic enzymes in 3D pellet cultures *in vitro*.** hMSCs, transduced with lentivirus to enable constitutive expression of full-length TSG-6 (or with control virus), were cultured as chondrocyte pellets for 14 days and then incubated with or without 10 ng/ml **(A)** IL-1α or **(B)** TNF for 24 h. *ADAMTS4*, *ADAMTS5* and *MMP13* gene expression levels were determined by qPCR and are presented as fold-change relative to the 'no virus, no cytokine' control. Data are shown for experiments performed in triplicate with hMSCs from Donors A (red circles) and C (black triangles), with mean values (n=6) indicated by horizontal lines. Data were analysed using two-way ANOVA with Tukey's *post hoc* test; ****$p$<0.0001 relative to 'control virus + cytokine'.

**Supplemental Figure S7. Link_TSG6 reduces OA-associated joint damage in the rat ACLTpMMx model.** OA was induced in the right knees of male Lewis rats by transection of the anterior cruciate ligament (ACLT) and a partial (30%) medial meniscectomy (pMMx). Link_TSG6 treatment (or vehicle) was administered *i.a.* at 7, 14 and 21 days post-surgery, at the doses indicated. At the 28-day end point, joint tissues were fixed, decalcified and 7 μm sections were stained with Safranin O/Fast Green or Alcian Blue (pH 2.5), prior to blinded histopathological scoring by two individuals across 7 parameters (each on a scale of 0 to 4; see Supplementary Table 3). Total sum scores for the medial tibiae are shown, with data presented as scatter plots for each treatment group (n=18-20); the mean scores ± SD are indicated by horizontal bars. A dose-dependent reduction in mean total score was seen and unpaired t tests showed a significant reduction in joint damage (*$p$<0.05) following treatment with the 120 μg dose of Link_TSG6 compared to the vehicle-treated control.

### SUPPLEMENTAL FIGURE S1

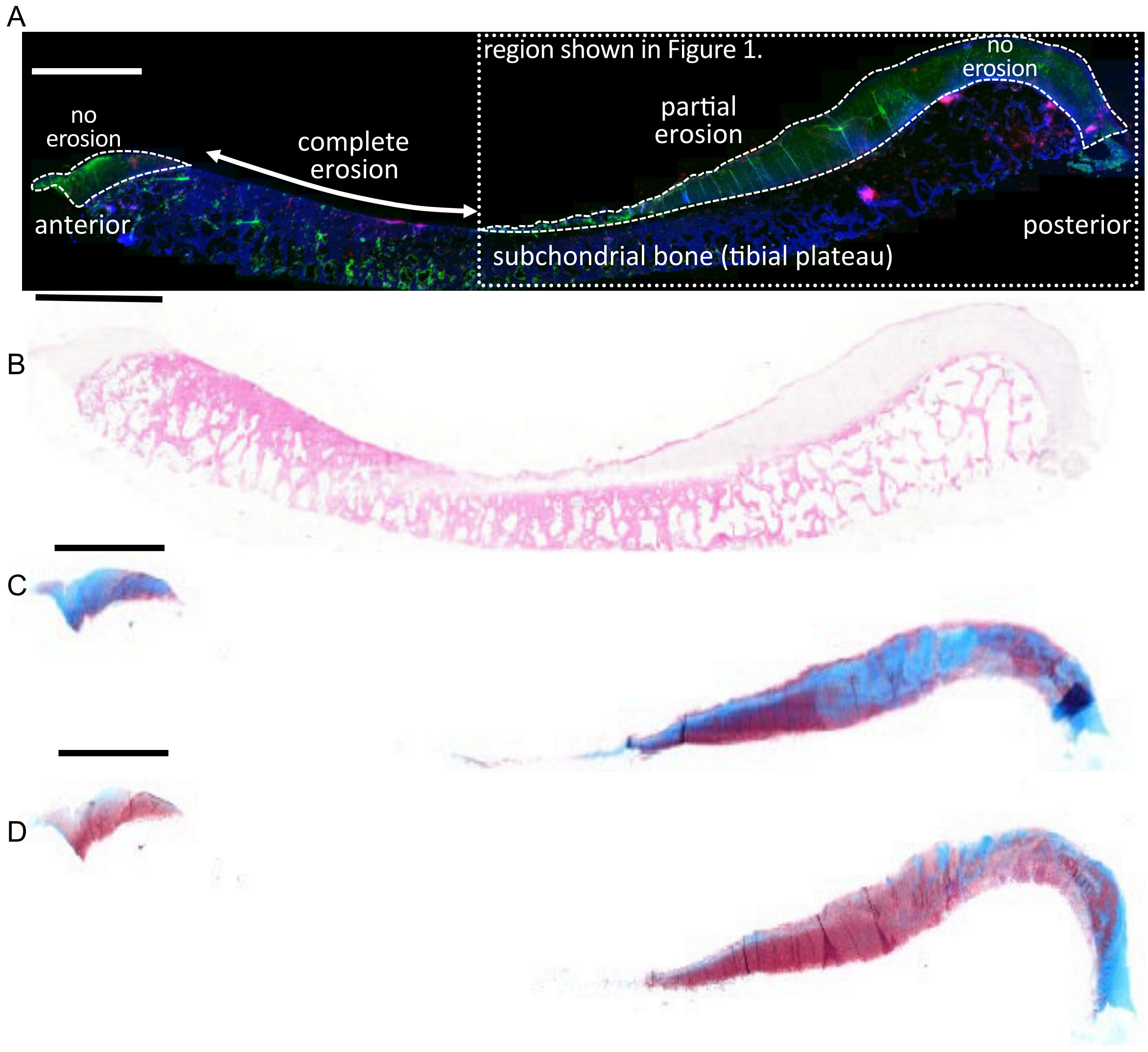

SUPPLEMENTAL FIGURE S2

A

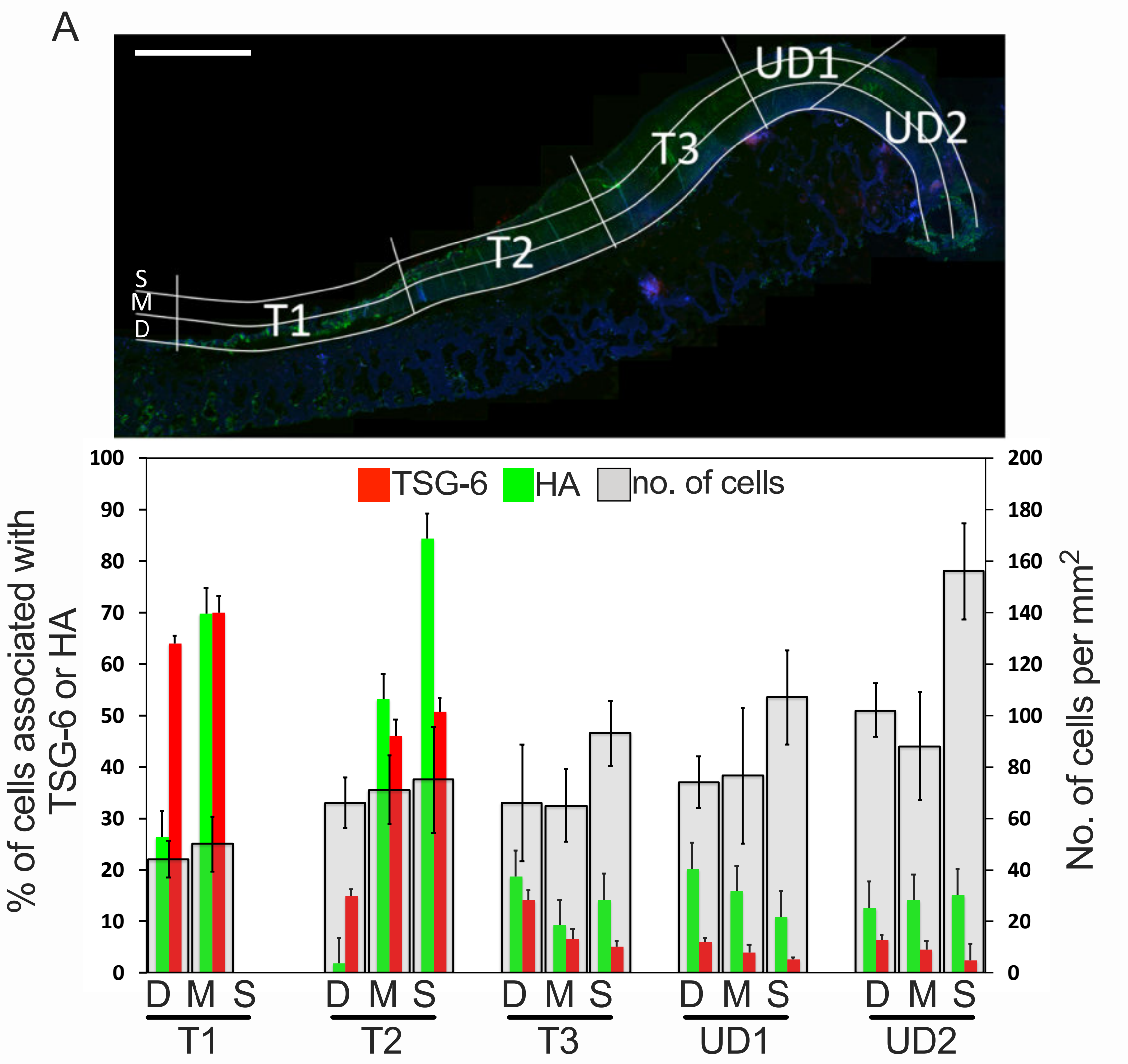


B

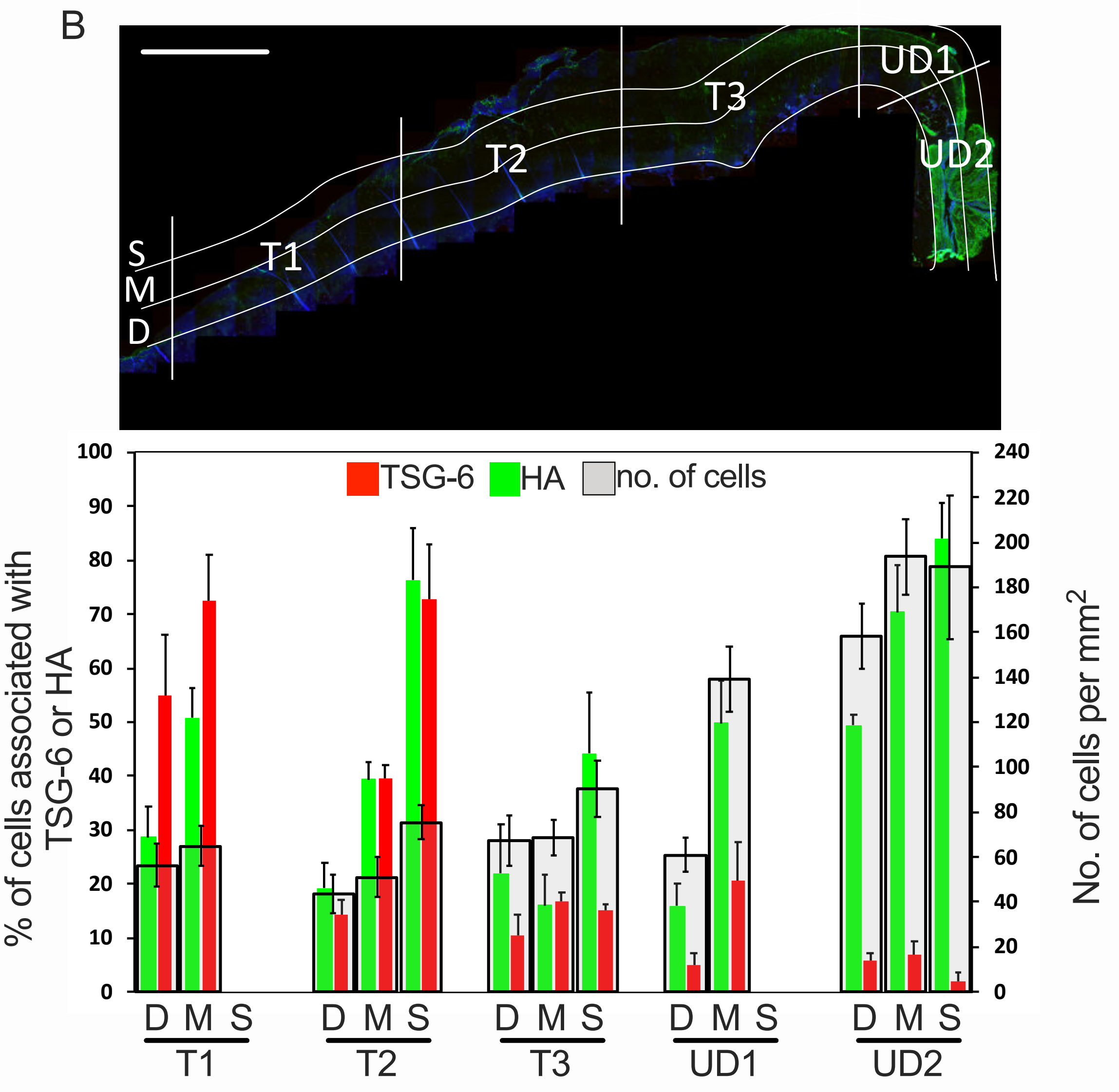

SUPPLEMENTAL FIGURE S3

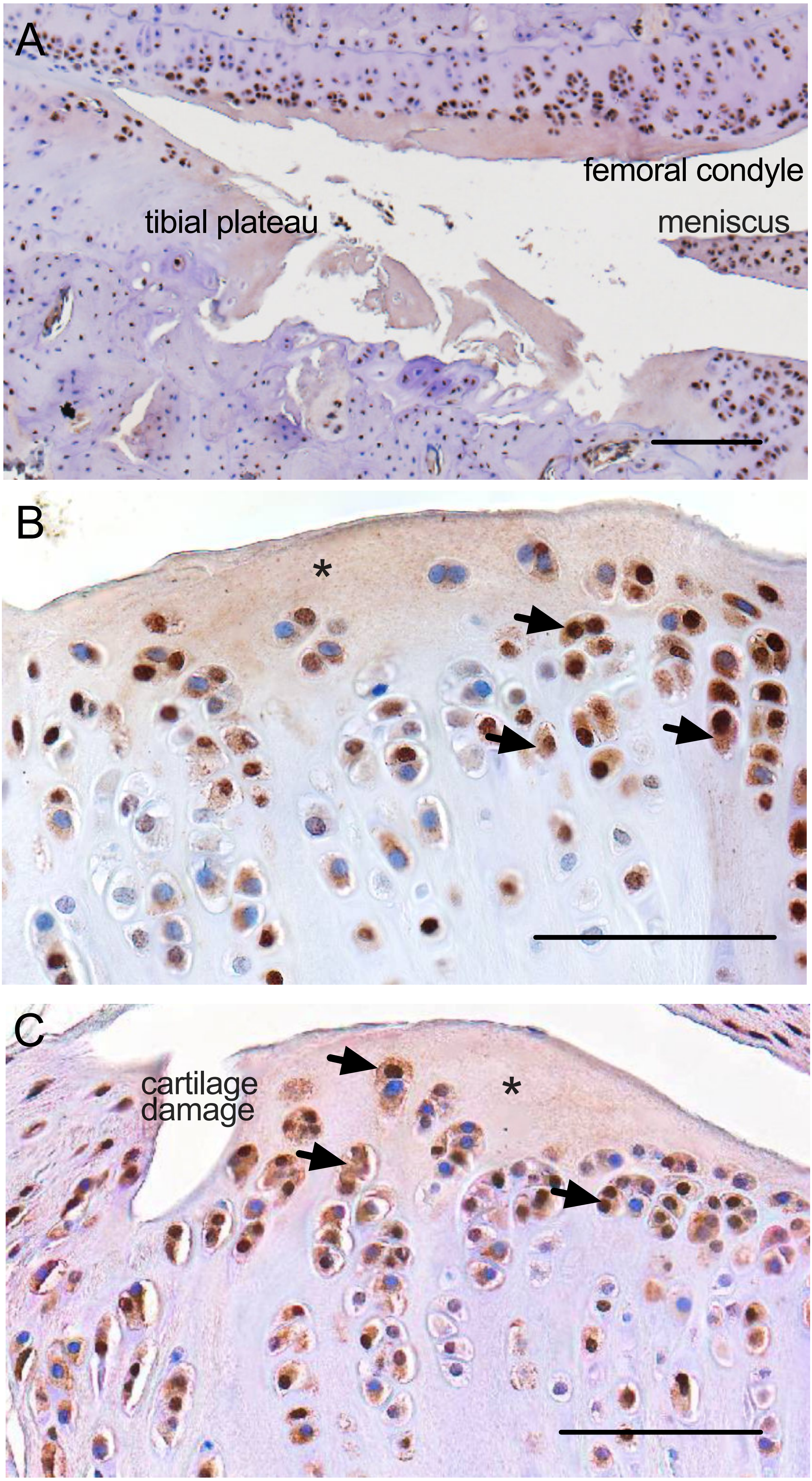

### SUPPLEMENTAL FIGURE S4

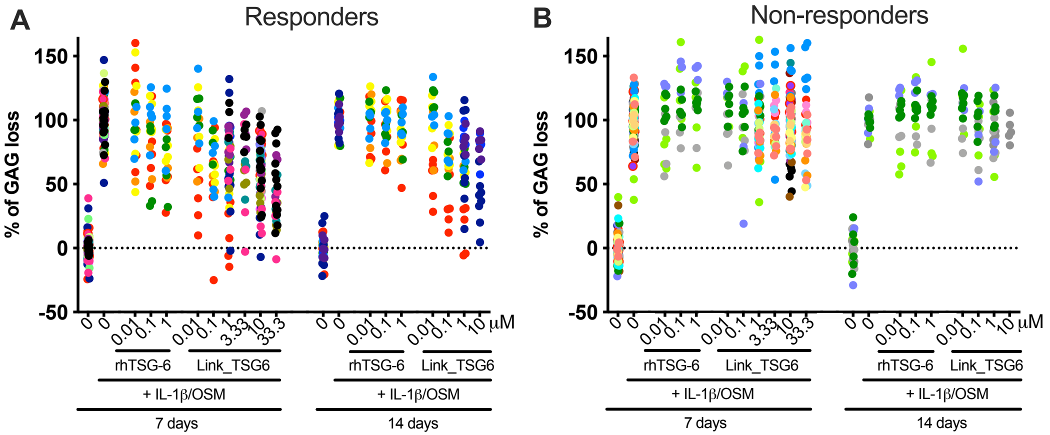

SUPPLEMENTAL FIGURE S5

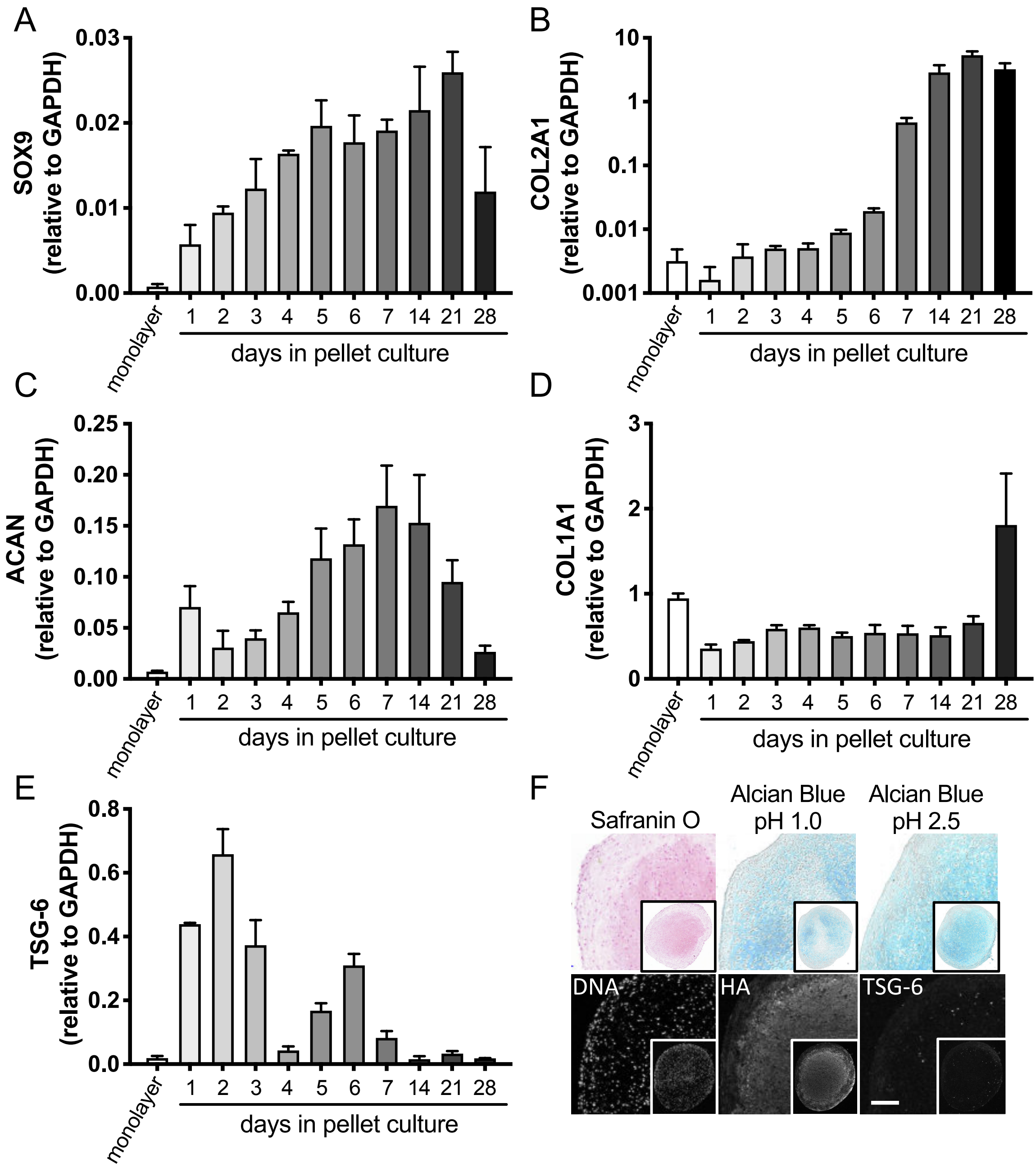

SUPPLEMENTAL FIGURE S6

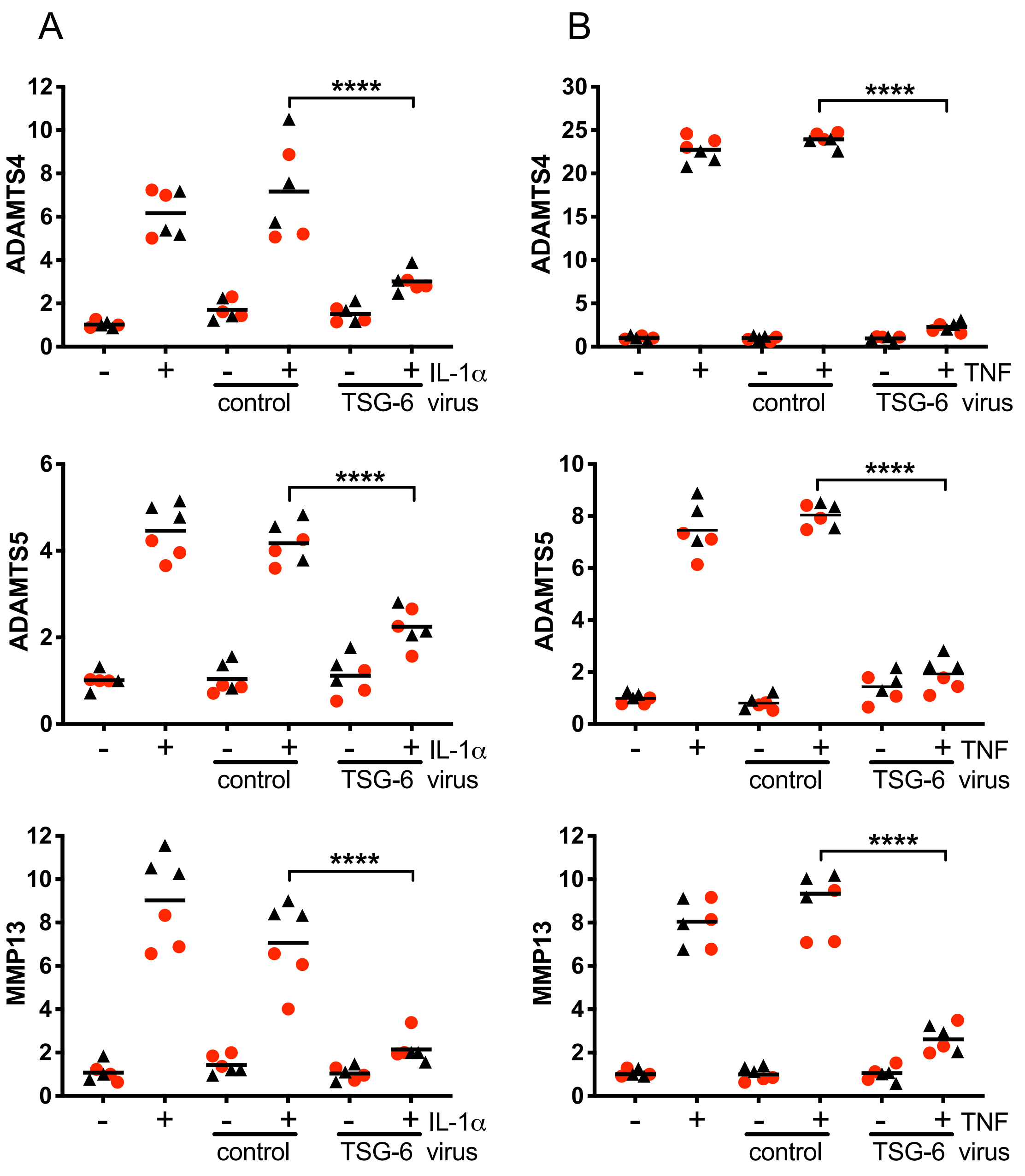

SUPPLEMENTAL FIGURE S7

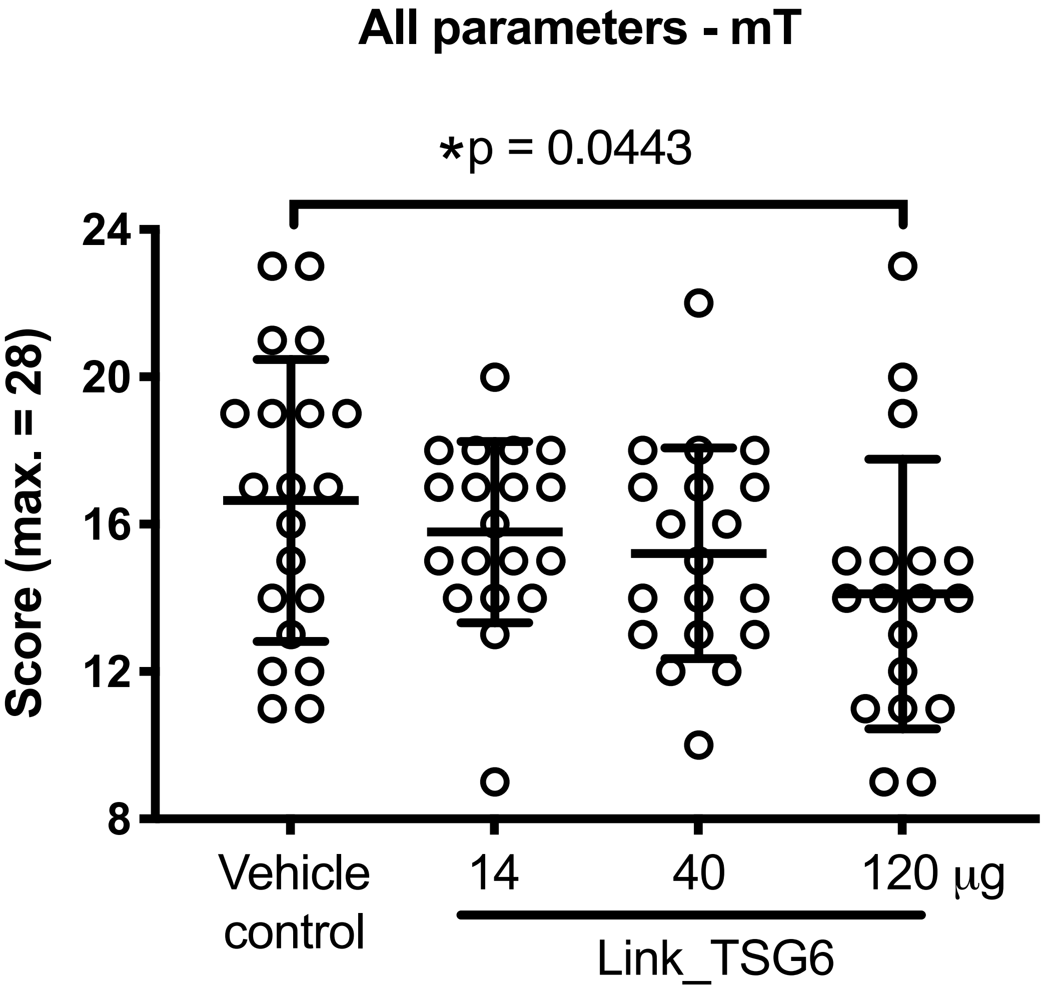
